# Identification of a distinctive gene signature in granulomatous myositis

**DOI:** 10.1101/2024.12.27.24319708

**Authors:** Iago Pinal-Fernandez, Nikolas Ruffer, Maria Casal-Dominguez, Katherine Pak, Felix Kleefeld, Corinna Preusse, Raphael Kirou, Travis B. Kinder, Stefania Del Orso, Faiza Naz, Shamima Islam, Gustavo Gutierrez-Cruz, Margherita Milone, Albert Selva-O’Callaghan, Jose C. Milisenda, Udo Schneider, Teerin Liewluck, Werner Stenzel, Andrew L. Mammen

## Abstract

**Objectives:** Granulomatous myositis (GM) is defined by focal collections of activated macrophages that fuse to form multinucleated cells that aggregate into granulomas within skeletal muscle. This study aimed to elucidate the pathophysiology of GM by defining its specific transcriptomic profile.

**Methods:** Bulk RNA sequencing was performed on 722 muscle biopsies, including 38 from patients with GM, other myopathies, and healthy comparators. Spatial transcriptomics and immunohistochemistry assays were used to identify the location of specific transcripts and proteins within the tissue.

**Results:** The transcriptomic signature of GM muscle biopsies was characterized by high levels of IFNγ, IFNγ-inducible genes, and proinflammatory cytokine genes including IL1B, TNF, and TGFB1. The expression levels of 1293 specifically upregulated and 256 specifically downregulated genes were highly correlated with transcriptomic markers of disease activity. Support vector machine models using this gene set identified GM with an AUC of 99.6% (99.0%-99.9%) and an accuracy of 98.6% (98.2%-98.9%). Expression of CHIT1, the most significantly upregulated gene, strongly correlated with disease severity and was detected at the RNA and protein level in granulomas and giant cells.

**Conclusions:** GM is transcriptomically characterized by a strong IFNγ signature and overexpression of proinflammatory cytokines, including IL1B, TNF, and TGFB1. Additionally, it exhibits a unique transcriptomic profile, including CHIT1, which correlates with disease activity.

## INTRODUCTION

Inflammatory myopathies (IM) include several distinct types of systemic autoimmune disease, including dermatomyositis (DM), the antisynthetase syndrome (ASyS), immune-mediated necrotizing myopathy (IMNM), and inclusion body myositis (IBM). Each type of IM is associated with unique muscle biopsy features, ranging from diffuse myofiber necrosis in IMNM to the invasion of myofibers by cytotoxic T cells in patients with IBM.[1] Granulomatous myositis (GM) is a form of IM defined histologically by the formation of non-caseating granulomas within muscle tissue.[2,3]

Transcriptomic analyses have provided important insights into the pathophysiological processes underlying the different types of IM.[4–8] For example, recent studies have demonstrated that in DM patients with anti-Mi2 autoantibodies, internalization of these autoantibodies into muscle cells disrupts the ability of Mi2 to repress a specific set of genes, leading to the aberrant expression of this gene set.[4,7] To date, the transcriptomic profile of muscle tissue from patients with GM has not been defined.

In the current study, we analyzed gene expression patterns in GM muscle biopsies and compared these with the transcriptomic profiles of muscle tissues from other types of IM, disease controls, and healthy controls. Having defined a GM-specific gene set, we then assessed its association with transcriptomic features of disease activity and also evaluated the utility of the GM-specific gene set to identify GM using various machine learning models.

## METHODS

### Patients

Muscle biopsies from myopathy patients were collected from Institutional Review Board (IRB)-approved longitudinal cohorts at the National Institutes of Health (Bethesda, MD), Johns Hopkins Myositis Center (Baltimore, MD), Vall d’Hebron Hospital (Barcelona), Clinic Hospital (Barcelona), Mayo Clinic (Rochester, MN), and Charité-Universitätsmedizin (Berlin). Patients were classified as IBM if they met Lloyd’s criteria.[9] Autoantibodies were detected by ELISA, immunoprecipitation of in vitro transcription and translation-generated proteins (IVTT-IP), EUROLINE myositis profile line blotting, or immunoprecipitation from ^35^S-methionine-labeled HeLa cell lysates. Patients testing positive for a myositis-specific autoantibody were classified based on their myositis-specific autoantibody according to the Casal and Pinal criteria[10]: those positive for anti-Jo1, anti-PL7, anti-PL12, anti-EJ or anti-OJ were classified as ASyS; those positive for anti-Mi2, anti-NXP2, anti-MDA5, anti-TIF1g or anti-SAE1 were classified as DM; and those with autoantibodies against SRP or HMGCR were classified as IMNM. Additionally, patients who met the 2017 ACR/EULAR classification criteria for DM[11] but were negative for dermatomyositis-specific autoantibodies were also classified as having DM.

### Ethics Approval and Consent

The study received IRB approval from the institutions involved. Written informed consent was obtained from all participants, and all methods were performed following relevant guidelines and regulations.

### RNA Sequencing

Bulk RNA sequencing (RNAseq) was conducted on frozen muscle biopsy specimens, following previously established protocols.[4–7,12] Muscle samples were immediately flash-frozen upon biopsy and stored at -80°C at each participating center. They were then transported on dry ice to the NIH, where they were uniformly processed. RNA extraction was performed using TRIzol. Libraries were prepared either using the NeoPrep system with the TruSeq Stranded mRNA Library Prep protocol (Illumina, San Diego, CA) or with the NEBNext Poly(A) mRNA Magnetic Isolation Module and Ultra™ II Directional RNA Library Prep Kit for Illumina (New England BioLabs, ref. #E7490 and #E7760).

### Pathology and immunohistochemistry

Muscle biopsy slides processed for clinical evaluation were stained with hematoxylin and eosin, along with relevant enzyme immunohistochemistry reactions on 8-μm cryostat sections, following international guidelines.[13] To confirm negative staining results, irrelevant antibodies were included as controls using the same antibody panel for validation.

### Statistical and Bioinformatic Analysis

RNA sequencing reads were demultiplexed using bcl2fastq 2.20.0 and preprocessed with fastp 0.23.4. Gene abundance was quantified with Salmon 1.5.2, and counts were normalized using the Trimmed Means of M values (TMM) method in edgeR 4.2.1 for graphical representation. Differential gene expression analysis was conducted with limma 3.60.6, applying Benjamini-Hochberg correction for multiple testing where appropriate. Pathway enrichment analysis was performed using a one-sided Fisher’s exact test.

To identify gene sets associated with each specific group, we calculated the intersection of differentially overexpressed genes (q-value < 0.01) between each target group and its comparator groups. Venn diagrams were used to visually represent these intersections.

To systematically analyze interleukins, chemokines, TNF superfamily members, and their receptors, we filtered the HGNC dataset to include these gene groups.

Diagnostic machine learning models were developed in Python using Scikit-Learn 1.5.2, with default parameters applied across models. Model performance was assessed through stratified 10-fold cross-validation.[14] To construct confidence intervals, we calculated the 2.5% and 97.5% percentiles of performance metrics from 1,000 iterations of the stratified 10-fold cross-validation process, each with varying random seeds.

Spatial transcriptomic data from sarcoid myopathy was retrieved from the publicly available dataset GSE243291.[15] The data was downloaded, normalized, and analyzed using Seurat version 5.1.0.

## RESULTS

### General Transcriptional Features of Granulomatous Muscle Biopsies

This study included 38 muscle biopsies from patients with GM, drawn from three different patient cohorts (Supplementary Table 1). Muscle biopsies from patients with GM had a marked upregulation of IFNγ and IFNγ-inducible genes, comparable in magnitude to those seen in muscle biopsies from patients with IBM (Figure 1, Supplementary Figures 1-3). IFNγ receptor genes were also notably upregulated to a greater extent than in other forms of inflammatory myopathies (Supplementary Figure 2). In contrast, IFNα and IFNβ genes were undetectable, though IFNα receptor 2 was significantly upregulated at levels similar to those observed in IBM. Type I IFN-inducible genes, such as ISG15 and MX1, were elevated at intermediate levels, lower than those seen in DM but similar to antisynthetase syndrome (Figure 1, Supplementary Figures 1-2).

**Figure 1.**
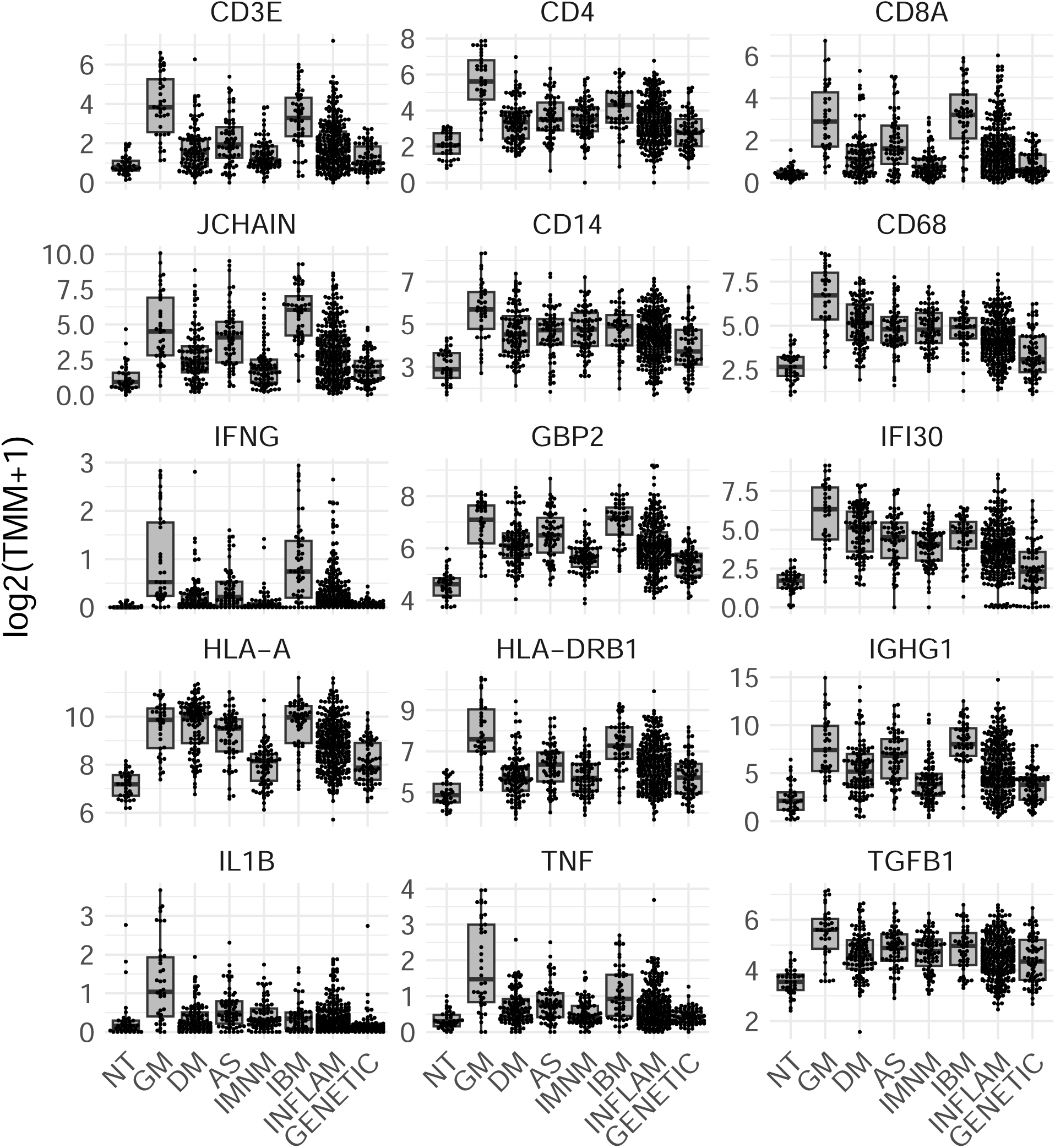
Expression of representative genes in granulomatous myositis compared to other myopathies. Each dot represents the gene expression value of a single patient. NT: histologically normal muscle biopsies; GM: granulomatous myositis; DM: dermatomyositis; AS: antisynthetase syndrome; IBM: inclusion body myositis; INFLAM: inflammatory myopathies; GENETIC: genetic myopathies.

A significant upregulation of T-cell markers was observed, with a relative predominance of CD4 over CD8 expression (Supplementary Figure 4). Additionally, B-cell markers (*MS4A1*) and plasma cell markers (*JCHAIN* and *SDC1*) were significantly elevated compared to most other inflammatory myopathy groups, reaching levels akin to those seen in IBM muscle biopsies (Supplementary Figure 4). Local production of various immunoglobulin isotypes was also elevated (Supplementary Figure 5). As expected, macrophage markers (CD14 and CD68) were upregulated at higher levels than in other inflammatory myopathies (Supplementary Figure 4). Levels of canonical endothelial cell markers (VWF) were not significantly different between GM and control biopsies, but venular endothelial cell markers (ACKR1) were elevated to levels seen in the other types of IM (Supplementary Figure 4).

Type I HLA expression was elevated in GM to levels comparable to that seen in IBM muscle biopsies. However, HLA type II expression (induced by IFNγ and produced by T-cells) was upregulated to higher levels in GM than in any other type of IM (Supplementary Figure 5).

The expression of genes serving as markers of type I myofibers (*MYH7*) were more markedly decreased than genes serving as markers for type II fibers (*MYH2* and *MYH1*) (Supplementary Figure 6). The genes encoding other structural proteins, such as ACTA1 and TTN, were reduced at levels similar to those in DM biopsies (Supplementary Figure 6). Mitochondrial markers were globally decreased in GM, reflecting a pattern seen in other forms of IM (Supplementary Figure 7).

A systematic analysis of interleukin gene expression revealed substantial upregulation of multiple proinflammatory interleukins, including IL1B, CXCL8 (IL8), IL10, IL26, and IL16 (Supplementary Figure 8). Complementary upregulation of interleukin receptors was also observed with IL1R, IL2R, IL17R, and IL10R being notably increased (Supplementary Figure 9). In addition, checkpoint ligands and their corresponding receptors were elevated (Supplementary Figure 10).

Similarly, genes encoding several chemokines, such as CCL17, CCL22, CCL18, CCL13, CCL16, XCL1, CCL3, and CCL4, were specifically upregulated in GM (Supplementary Figure 11). Chemokine receptor genes including CXCR6 (ligand of CCL16), XCR1 (ligand of XCL1), CCR5 (ligand of CCL3 and CCL4), and CCR2 (ligand of CCL2, CCL7, CCL8, and CCL13) were also prominently upregulated (Supplementary Figure 12).

The genes for multiple TNF family molecules, including TNF, CD40LG, TNFSF13, LTA, TNFSF14, FASLG, and CD70, showed specific upregulation in granulomatous myositis (Supplementary Figure 13). Similar to interleukins, the genes encoding their receptors were concurrently elevated, with increases in TNFRSF1B (receptor for TNF), CD40 (receptor for CD40LG), TNFRSF14 (ligand of TNFSF14), CD27 (ligand of CD70), and FAS (receptor of FASLG) (Supplementary Figure 14).

Finally, TGFB1 and TGFB3 were upregulated in GM, with TGFB1 particularly overexpressed compared to other forms of myopathy. The receptor for TGFB2 was also elevated in GM (Supplementary Figure 15).

Spatial transcriptomics revealed that several key findings are specifically localized to the non-caseating granulomas, including CD4+ T-cell-dominated infiltrates, the presence of plasma cells with local immunoglobulin production, the expression of IFNγ and its inducible genes, and the production of IL1B, TNF, and TGFB1 (Supplementary Figure 16).

### A Specific Gene Set is Associated with Granulomatous Myositis

To identify genes specifically associated with GM, we calculated the intersection of differentially expressed genes (q-value cutoff < 0.01) between biopsies from patients with GM and each comparator group (Figure 2). This analysis revealed 1293 genes that were specifically upregulated in GM (Figure 3; Table 1) and 256 genes that were specifically downregulated in GM (Supplementary Figure 17; Table 1). The upregulated genes exhibited considerably more significant differential expression compared to the downregulated genes (Table 1). As anticipated, these genes were predominantly expressed by macrophages and were generally associated with terminally differentiated, highly active macrophage populations located in the granulomas of the muscle (Figure 4).

**Figure 2.**
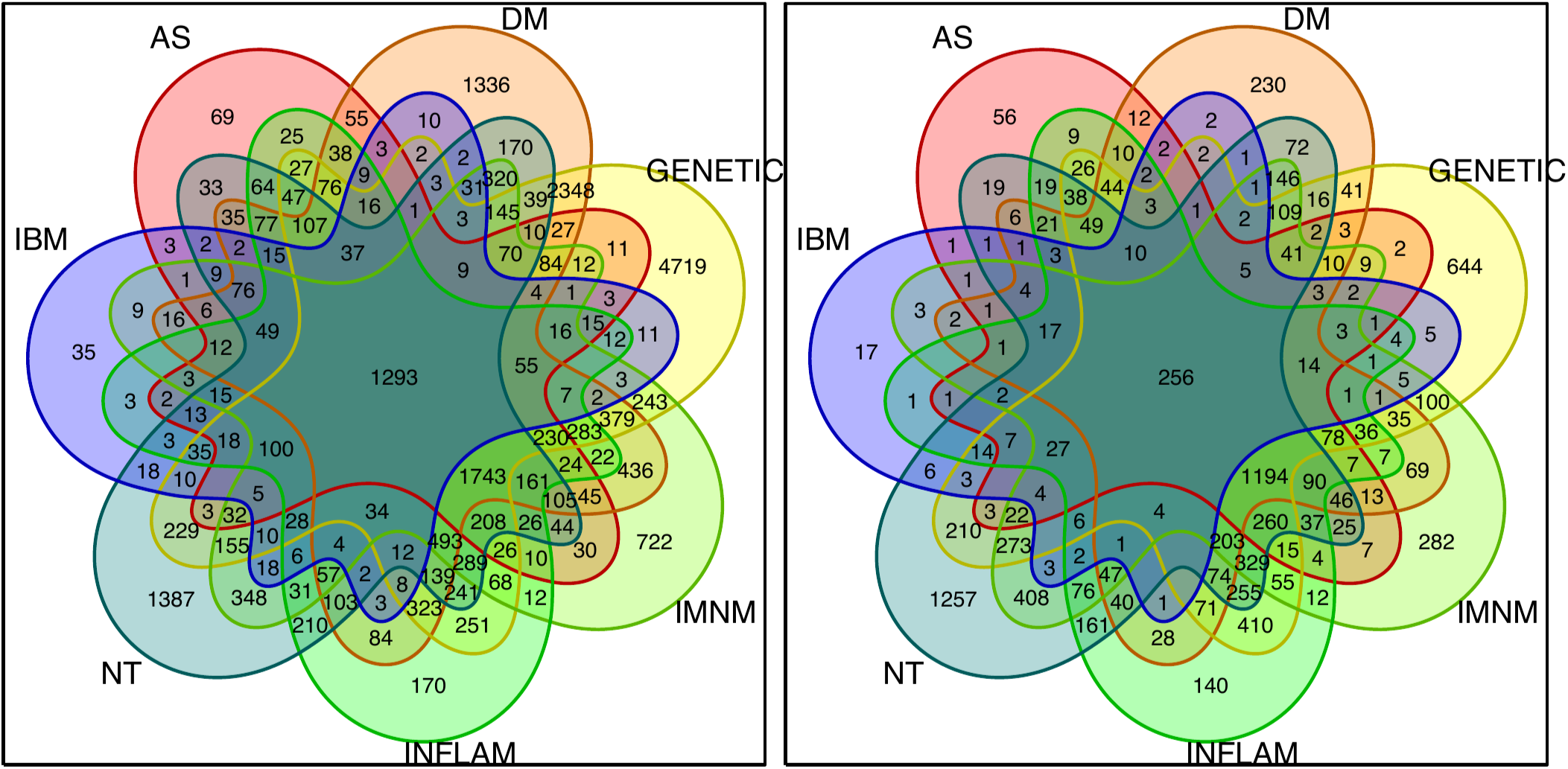
Venn diagrams showing the number of genes that were differentially (q-value < 0.01) overexpressed (left panel) and underexpressed (right panel) in patients with granulomatous myositis compared to each of the other myositis groups, other myopathies, and normal muscle biopsies. NT: histologically normal muscle biopsies; DM: dermatomyositis; AS: antisynthetase syndrome; IBM: inclusion body myositis; INFLAM: inflammatory myopathies; GENETIC: genetic myopathies.

**Figure 3.**
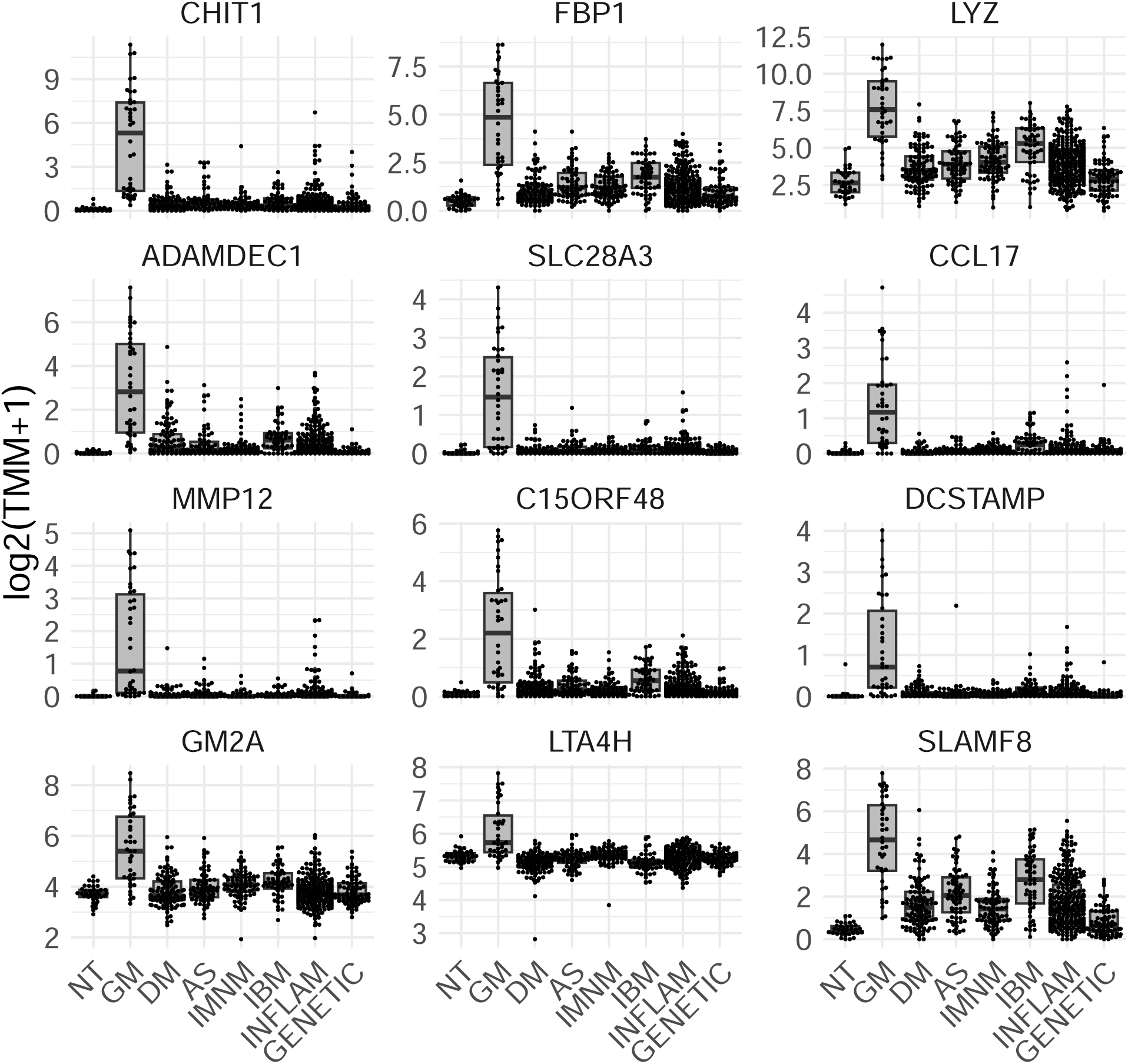
Expression of top 12 specifically overexpressed genes in granulomatous myositis compared to other myopathies. Each dot represents the gene expression value of a single patient. NT: histologically normal muscle biopsies; GM: granulomatous myositis; DM: dermatomyositis; AS: antisynthetase syndrome; IBM: inclusion body myositis; INFLAM: inflammatory myopathies; GENETIC: genetic myopathies.

**Table 1.** Differential expression of genes specifically overexpressed (left panel) and underexpressed (right panel) in granulomatous myositis patients compared to other muscle biopsy samples in the study. Pos: position in the differential expression table; logFC: log2 fold-change; adj.P.Val: adjusted p-value.

| Specifically overexpressed |  |  |  | Specifically underexpressed |  |  |  |
| --- | --- | --- | --- | --- | --- | --- | --- |
| Gene | Pos | logFC | adj.P.Val | Gene | Pos | logFC | adj.P.Val |
| CHIT1 | 1 | 6.3 | 1.7e-111 | GADD45GIP1 | 371 | -1.2 | 2.8e-22 |
| FBP1 | 2 | 4.2 | 1.6e-78 | MRFAP1 | 431 | -0.7 | 5.0e-21 |
| LYZ | 3 | 4.1 | 1.8e-74 | NDUFAF8 | 494 | -1.3 | 4.2e-20 |
| ADAMDEC1 | 4 | 5.4 | 1.0e-68 | TXNL4A | 571 | -0.8 | 6.4e-19 |
| SLC28A3 | 5 | 4.4 | 2.5e-65 | TAF13 | 645 | -0.9 | 4.9e-18 |
| CCL17 | 6 | 4.5 | 2.5e-65 | DYNLL2 | 739 | -1.0 | 5.1e-17 |
| MMP12 | 7 | 4.9 | 1.0e-63 | STUB1 | 764 | -0.9 | 8.9e-17 |
| C15ORF48 | 9 | 4.0 | 8.3e-58 | GNA11 | 765 | -0.8 | 8.9e-17 |
| DCSTAMP | 10 | 4.2 | 7.5e-53 | ANAPC11 | 772 | -1.0 | 1.0e-16 |
| GM2A | 11 | 1.7 | 1.8e-51 | RABL6 | 789 | -1.0 | 1.5e-16 |
| LTA4H | 12 | 0.9 | 1.8e-51 | MRPL12 | 791 | -1.1 | 1.6e-16 |
| SLAMF8 | 13 | 3.8 | 7.8e-50 | HNRNPM | 797 | -0.7 | 2.0e-16 |

Both the upregulated (Figure 5) and downregulated gene sets (Supplementary Figure 18) were strongly correlated with markers of disease activity in muscle biopsies. These markers included macrophage markers, type I and type II interferon-inducible genes, T-cell markers, muscle differentiation markers such as PAX7, as well as mitochondrial and mature muscle markers, which serve as transcriptomic proxies for muscle atrophy.

**Figure 5.**
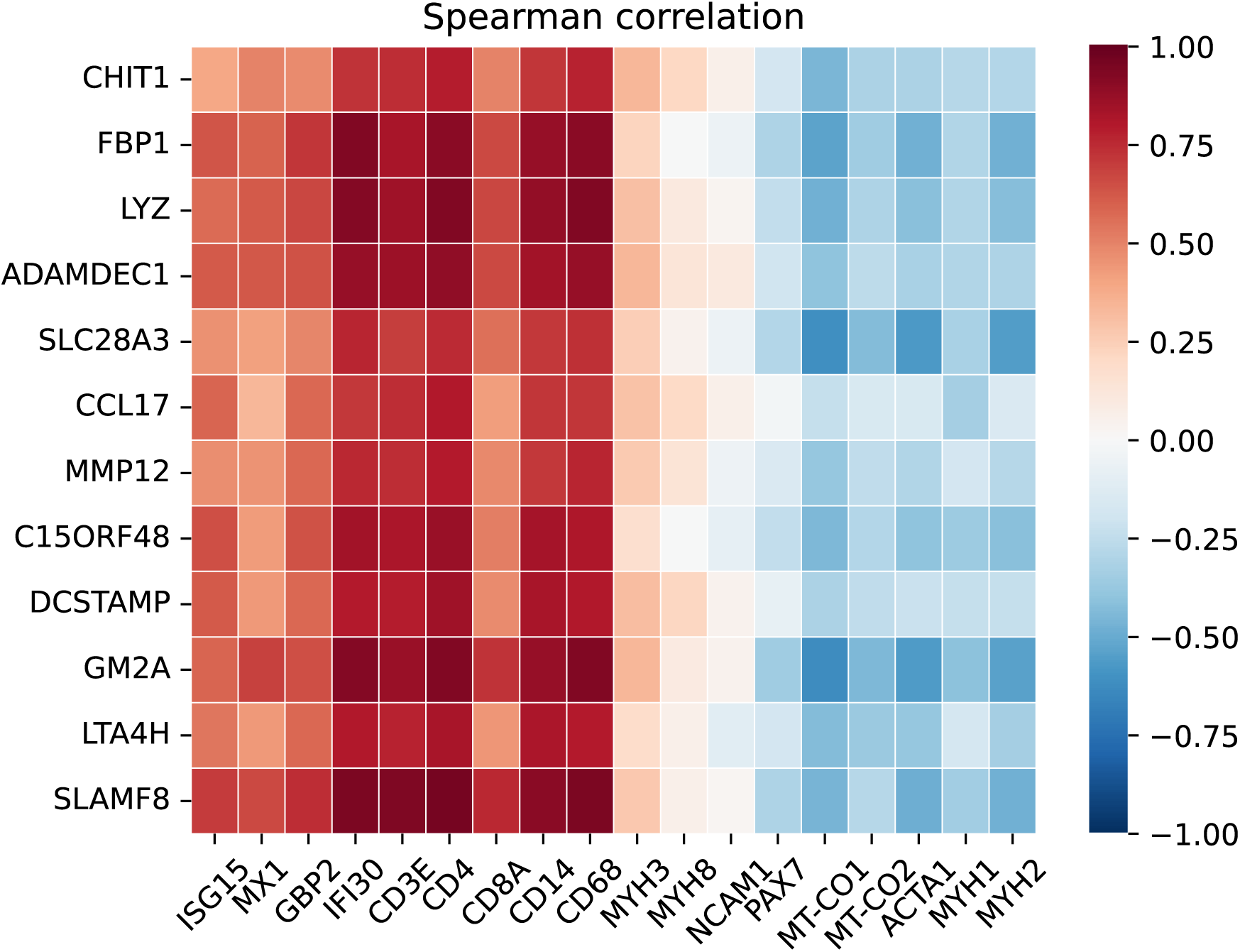
Correlation of the top 12 specifically overexpressed genes in granulomatous myositis with transcriptomic markers of disease activity, including type 1 interferon-inducible genes (ISG15, MX1), type 2 interferon-inducible genes (GBP2, IFI30), T-cell markers (CD3E, CD4, CD8), macrophage markers (CD14, CD68), markers of muscle differentiation (MYH3, MYH8, NCAM1, PAX7), mitochondrial markers (MT-CO1, MT-CO2), and mature muscle structural proteins (ACTA1, MYH1, MYH2).

Spatial transcriptomic analysis of publicly available GM data[15] confirmed that the most overexpressed genes we identified were specifically localized within the noncaseating granulomas (Figure 6).

**Figure 6.**
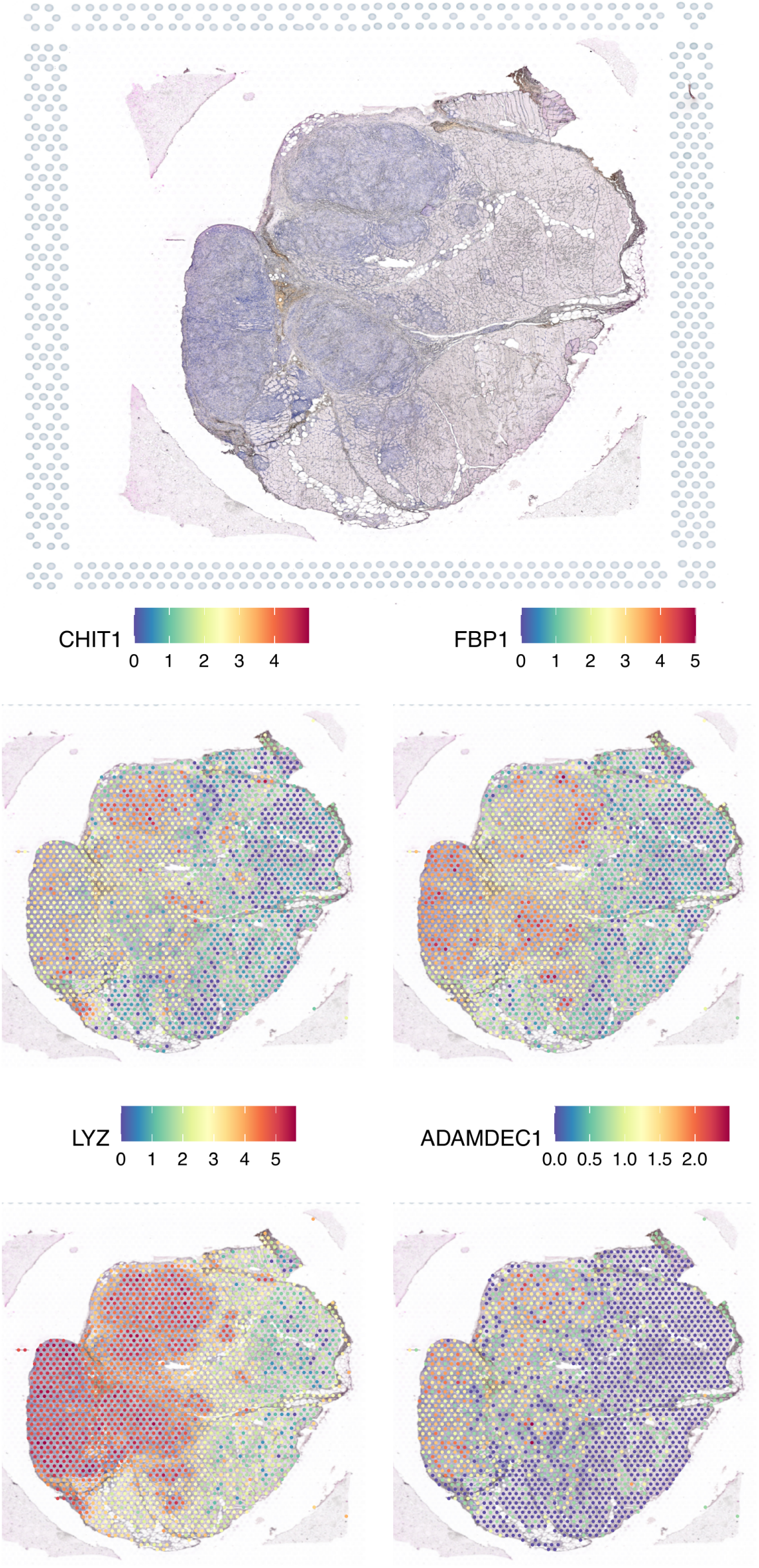
Distribution of the expression levels of the top four genes specifically overexpressed in granulomatous myositis, localized within the granulomas.

Among these, CHIT1 was the most significantly upregulated gene in patients with GM. This upregulation was also observed at the protein level in noncaseating granulomas and giant cells (Figure 7).

**Figure 7.**
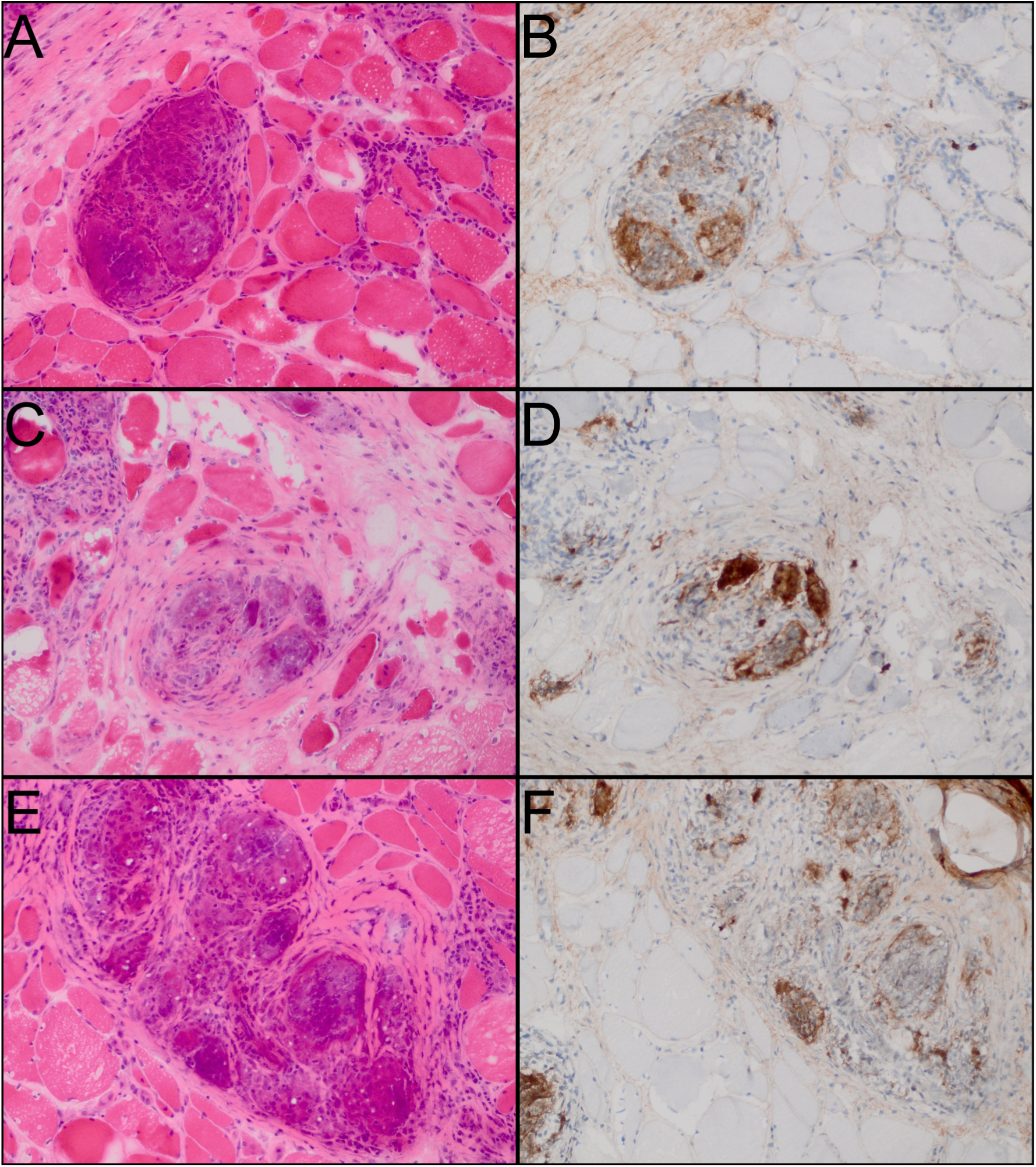
Immunohistochemistry for CHIT1 in muscle biopsies from patients with granulomatous myositis reveals intense staining in non-caseating granulomas and giant cells.

Transcriptomic profiles across the various cohorts showed no significant differences (Supplementary Figure 19).

### Performance of Machine Learning Models Derived from Specific Gene Sets

Several machine learning models were developed using both upregulated and downregulated gene sets. Consistent with previous studies,[5] the support vector machine (SVM) model emerged as the top performer in terms of accuracy, achieving an area under the ROC curve (AUC) of 99.6% (99.0%-99.9%) and an accuracy of 98.6% (98.2%-98.9%), establishing it as an excellent predictive classifier (Supplementary Table 2).

## DISCUSSION

In this study, we define the specific transcriptomic features of GM, revealing a distinct activation of the IFNγ pathway compared to other myopathies, as well as an increase in multiple proinflammatory cytokines, including IL1B, TNF, and TGFB1. Additionally, we identified a specific gene set that not only distinguishes GM from other conditions but also strongly correlates with disease activity. Using this gene set, we developed classification models with excellent predictive accuracy based on transcriptomic data.

Several of the inflammatory pathways identified, including activation of the IFN-γ pathway and upregulation of proinflammatory cytokines (e.g., IL1B, TNF, and IFNG), have also been observed in other affected tissues in patients with sarcoidosis, where blocking these pathways has demonstrated clinical efficacy.[16–19] For instance, TNF production by alveolar macrophages is well-documented,[16,17] and TNF inhibitors have proven effective in managing various manifestations of the disease.[20] Similarly, IL1B has been implicated in sarcoidosis,[17–19] and IL1 blockade with anakinra has shown promising results in cardiac sarcoidosis, including reductions in C-reactive protein (CRP) levels.[21] Additionally, case reports and a small clinical trial suggest that JAK/STAT inhibitors may also be effective in sarcoidosis, underscoring the potential of targeting this pathway in GM as well.[22,23] The identification of these pathways in muscle tissue from our patients suggests that similar therapeutic strategies could offer potential benefits in GM, and warrants further investigation into targeting additional pathways we have identified as altered in this condition.

Our study identified a gene set that accurately differentiates muscle biopsies from GM patients from other forms of myopathy, suggesting that minimally invasive needle biopsies may be sufficient for GM diagnosis, enabling earlier detection and treatment. This gene expression profile was also strongly associated with disease activity, indicating these markers could be valuable for monitoring disease progression. Notably, CHIT1 was the most significantly overexpressed gene, consistent with reports of its elevation in the sera and lymph nodes of sarcoidosis patients, and its pharmacological blockade is currently being evaluated in clinical trials for active pulmonary sarcoidosis (NCT06205121).[24] These findings suggest that the gene expression effects observed in muscle biopsies may extend to other sarcoidosis-affected tissues, broadening the potential impact of this research.

The etiology of granulomatous myositis remains unclear. The transcriptomic profile identified here sets the stage for targeted studies to explore whether novel autoantibodies or somatic mutations contribute to the observed biological effects.

This study has some limitations. First, we did not include cases with infectious or genetic causes of granulomatous muscle diseases, as these samples were not available at the time of the study. Second, except for CHIT1, which we examined at both the RNA and protein levels, the other markers were identified only at the RNA level.

In conclusion, this study systematically defines the transcriptomic characteristics and specific gene expression profile of granulomatous myositis. These findings not only provide insights into the disease’s pathogenesis but also offer reliable markers for diagnosing GM with high accuracy and monitoring disease activity, potentially guiding future therapeutic strategies.

## Data Availability

All data produced in the present study are available upon reasonable request to the authors

## Contributorship

All authors contributed to the development of the manuscript, including interpretation of results, substantive review of drafts, and approval of the final draft for submission. IPF and ALM are the guarantors.

## Acknowledgments

This study was funded, in part, by the Intramural Research Program of the National Institute of Arthritis and Musculoskeletal and Skin Diseases, National Institutes of Health.

## Acknowledgments

To Julie Thompson for her invaluable help maintaining the NIH Natural History Protocol. To the NIAMS Sequencing Core and its members.

## Ethical approval information

All biopsies were from subjects enrolled in institutional review board (IRB)-approved longitudinal cohorts in their respective centers.

## Patient and public involvement

Patients and/or the public were not involved in the design, conduct, reporting, or dissemination plans of this research.

## Data sharing statement

Any anonymized data not published within the article will be shared by request from any qualified investigator.

## REFERENCES

1 Selva-O’Callaghan A, Pinal-Fernandez I, Trallero-Araguás E, et al. Classification and management of adult inflammatory myopathies. Lancet Neurol. 2018;17:816–28. doi: 10.1016/s1474-4422(18)30254-0

2 Lequain H, Streichenberger N, Gallay L, et al. Granulomatous myositis: characteristics and outcome from a monocentric retrospective cohort study. Neuromuscular Disord. 2024;42:5–13. doi: 10.1016/j.nmd.2024.06.007

3 Chompoopong P, Skolka MP, Ernste FC, et al. Symptomatic myopathies in sarcoidosis: disease spectrum and myxovirus resistance protein A expression. Rheumatology. 2022;62:2556–62. doi: 10.1093/rheumatology/keac668

4 Pinal-Fernandez I, Muñoz-Braceras S, Casal-Dominguez M, et al. Pathological autoantibody internalisation in myositis. Ann Rheum Dis. Published Online First: 20 June 2024. doi: 10.1136/ard-2024-225773

5 Pinal-Fernandez I, Casal-Dominguez M, Derfoul A, et al. Machine learning algorithms reveal unique gene expression profiles in muscle biopsies from patients with different types of myositis. Ann Rheum Dis. 2020;79:1234–42. doi: 10.1136/annrheumdis-2019-216599

6 Pinal-Fernandez I, Casal-Dominguez M, Derfoul A, et al. Identification of distinctive interferon gene signatures in different types of myositis. Neurology. 2019;93. doi: 10.1212/wnl.0000000000008128

7 Pinal-Fernandez I, Milisenda JC, Pak K, et al. Transcriptional derepression of CHD4/NuRD-regulated genes in the muscle of patients with dermatomyositis and anti-Mi2 autoantibodies. Ann Rheum Dis. 2023;82:1091–7. doi: 10.1136/ard-2023-223873

8 Greenberg SA, Pinkus JL, Pinkus GS, et al. Interferon-α/β–mediated innate immune mechanisms in dermatomyositis. Ann Neurol. 2005;57:664–78. doi: 10.1002/ana.20464

9 Lloyd TE, Mammen AL, Amato AA, et al. Evaluation and construction of diagnostic criteria for inclusion body myositis. Neurology. 2014;83:426–33. doi: 10.1212/wnl.0000000000000642

10 Casal-Dominguez M, Pinal-Fernandez I, Pak K, et al. Performance of the 2017 European Alliance of Associations for Rheumatology/American College of Rheumatology Classification Criteria for Idiopathic Inflammatory Myopathies in Patients With Myositis-Specific Autoantibodies. A&R. 2022;74:508–17. doi: 10.1002/art.41964

11 Lundberg IE, Tjärnlund A, Bottai M, et al. 2017 European League Against Rheumatism/American College of Rheumatology classification criteria for adult and juvenile idiopathic inflammatory myopathies and their major subgroups. Annals of the Rheumatic Diseases. 2017;76:1955–64. doi: 10.1136/annrheumdis-2017-211468

12 Amici DR, Pinal-Fernandez I, Christopher-Stine L, et al. A network of core and subtype-specific gene expression programs in myositis. Acta Neuropathol. 2021;142:887–98. doi: 10.1007/s00401-021-02365-5

13 Dubowitz V, Sewry C, Oldfors A. Muscle biopsy: Apractical approach. Muscle Biopsy. 2021;vii.

14 Kohavi R. A study of cross-validation and bootstrap for accuracy estimation and model selection. 1995;1137–43.

15 Lequain H, Dégletagne C, Streichenberger N, et al. Spatial transcriptomics reveals signatures of histopathological changes in muscular sarcoidosis. Cells. 2023;12. doi: 10.3390/cells12232747

16 Bachwich PR, Lynch JP, Larrick J, et al. Tumor necrosis factor production by human sarcoid alveolar macrophages. The American journal of pathology. 1986;125:421–5.

17 Steffen M, Petersen J, Oldigs M, et al. Increased secretion of tumor necrosis factor-alpha, interleukin-1-beta, and interleukin-6 by alveolar macrophages from patients with sarcoidosis. Journal of Allergy and Clinical Immunology. 1993;91:939–49. doi: 10.1016/0091-6749(93)90352-g

18 Devergne O, Emilie D, Peuchmaur M, et al. Production of cytokines in sarcoid lymph nodes: Preferential expression of interleukin-1β and interferon-γ genes. Human Pathology. 1992;23:317–23. doi: 10.1016/0046-8177(92)90114-i

19 Hunninghake GW. Release of interleukin-1 by alveolar macrophages of patients with active pulmonary sarcoidosis. The American review of respiratory disease. 1984;129:569–72.

20 Adler BL, Wang CJ, Bui T-L, et al. Anti-tumor necrosis factor agents in sarcoidosis: A systematic review of efficacy and safety. Seminars in arthritis and rheumatism. 2019;48:1093–104. doi: 10.1016/j.semarthrit.2018.10.005

21 Kron J, Crawford T, Mihalick V, et al. Interleukin-1 blockade in cardiac sarcoidosis: study design of the multimodality assessment of granulomas in cardiac sarcoidosis: Anakinra Randomized Trial (MAGiC-ART). Journal of Translational Medicine. 2021;19. doi: 10.1186/s12967-021-03130-8

22 Damsky W, Thakral D, Emeagwali N, et al. Tofacitinib treatment and molecular analysis of cutaneous sarcoidosis. New Engl J Med. 2018;379:2540–6. doi: 10.1056/NEJMoa1805958

23 Damsky W, Wang A, Kim DJ, et al. Inhibition of type 1 immunity with tofacitinib is associated with marked improvement in longstanding sarcoidosis. Nat Commun. 2022;13:3140. doi: 10.1038/s41467-022-30615-x

24 Dymek B, Sklepkiewicz P, Mlacki M, et al. Pharmacological inhibition of chitotriosidase (CHIT1) as a novel therapeutic approach for sarcoidosis. Journal of Inflammation Research. 2022;15:5621–34. doi: 10.2147/JIR.S378357

