## Supplementary Figures for "Identification of a distinctive gene signature in granulomatous myositis"

**Supplementary Figure 1.** Expression of interferon genes in granulomatous myositis compared to other myopathies. Each dot represents the gene expression value from a single patient. NT: histologically normal muscle biopsies; GM: granulomatous myositis; DM: dermatomyositis; AS: antisynthetase syndrome; IBM: inclusion body myositis; INFLAM: inflammatory myopathies; GENETIC: genetic myopathies.

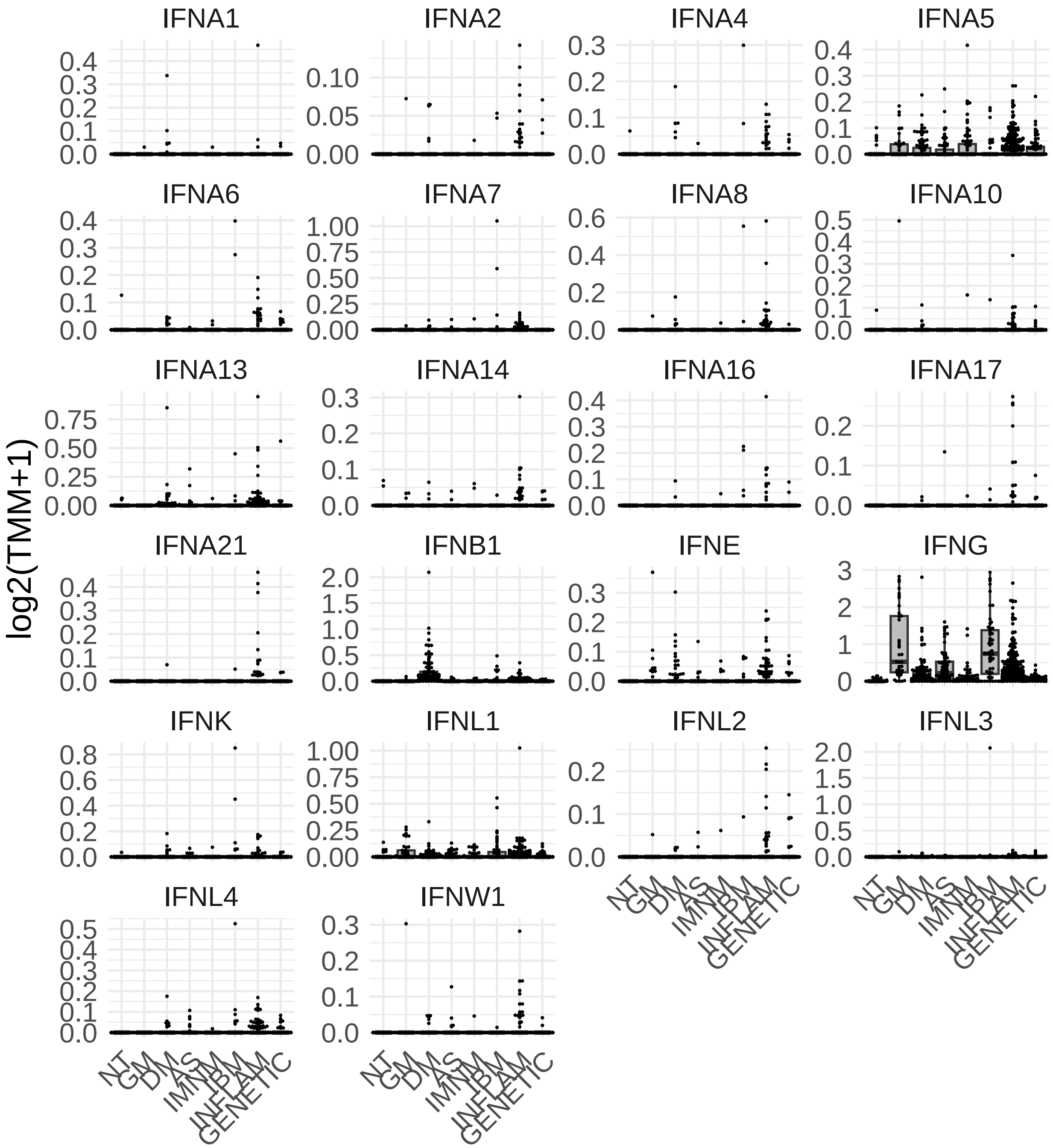

**Supplementary Figure 2.** Expression of type 1 and type 2 interferon-inducible genes and interferon receptors in granulomatous myositis compared to other myopathies. Each dot represents the gene expression value from a single patient. NT: histologically normal muscle biopsies; GM: granulomatous myositis; DM: dermatomyositis; AS: antisynthetase syndrome; IBM: inclusion body myositis; INFLAM: inflammatory myopathies; GENETIC: genetic myopathies.

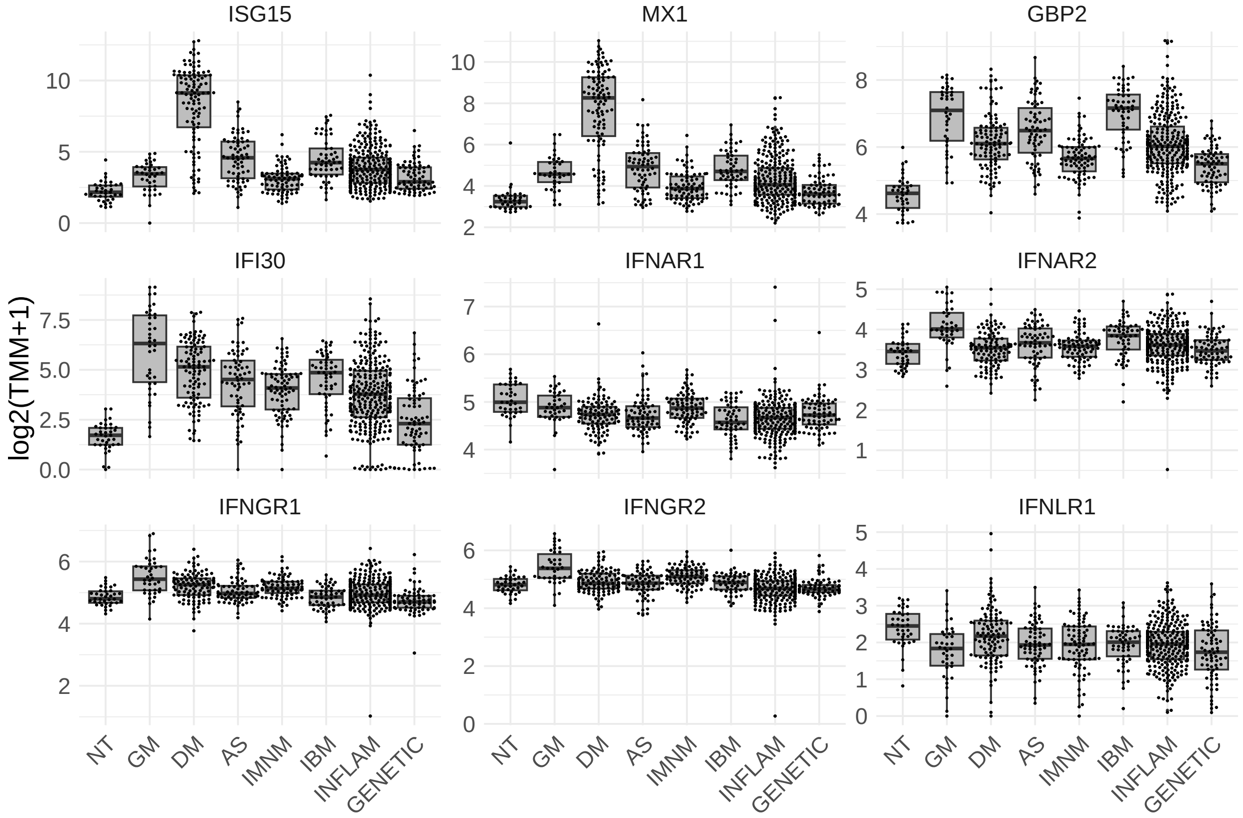

**Supplementary Figure 3.** Gene Set Enrichment Analysis of the hallmark gene sets from the Human Molecular Signatures Database in patients with granulomatous myositis compared to normal muscle. NES: normalized enrichment score, p-adjust: adjusted p-value

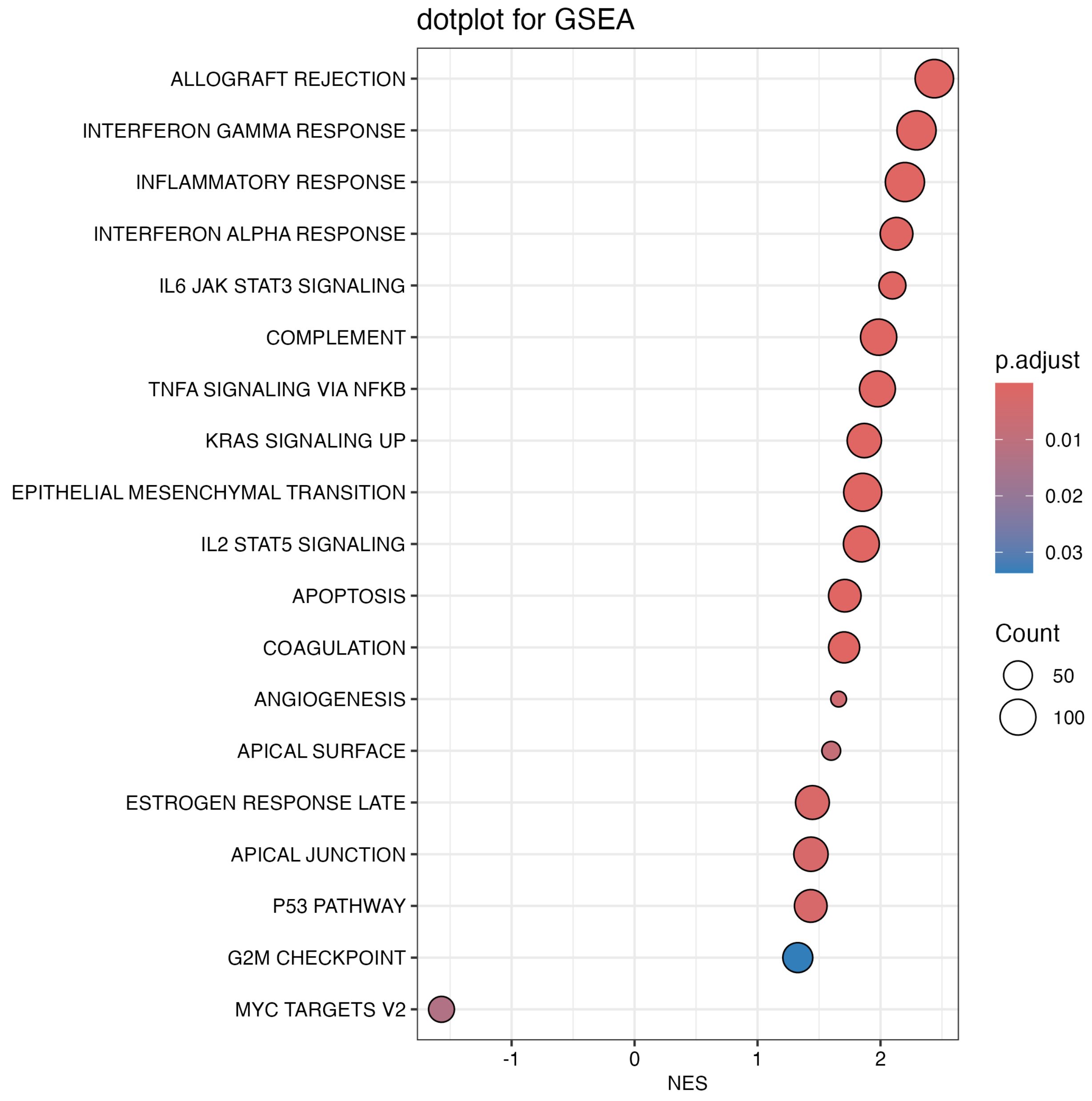

**Supplementary Figure 4.** Expression of transcriptomic markers associated with various cell types in granulomatous myositis compared to other myopathies. Markers include T-cell markers (CD3E, CD4, CD8A), B-cell markers (CD19, MS4A1), plasma cell markers (SDC1, JCHAIN), neutrophil markers (ELANE), macrophage markers (CD14, CD68), and endothelial cell markers (VWF, ACKR1). Each dot represents the gene expression value from a single patient. NT: histologically normal muscle biopsies; GM: granulomatous myositis; DM: dermatomyositis; AS: antisynthetase syndrome; IBM: inclusion body myositis; INFLAM: inflammatory myopathies; GENETIC: genetic myopathies.

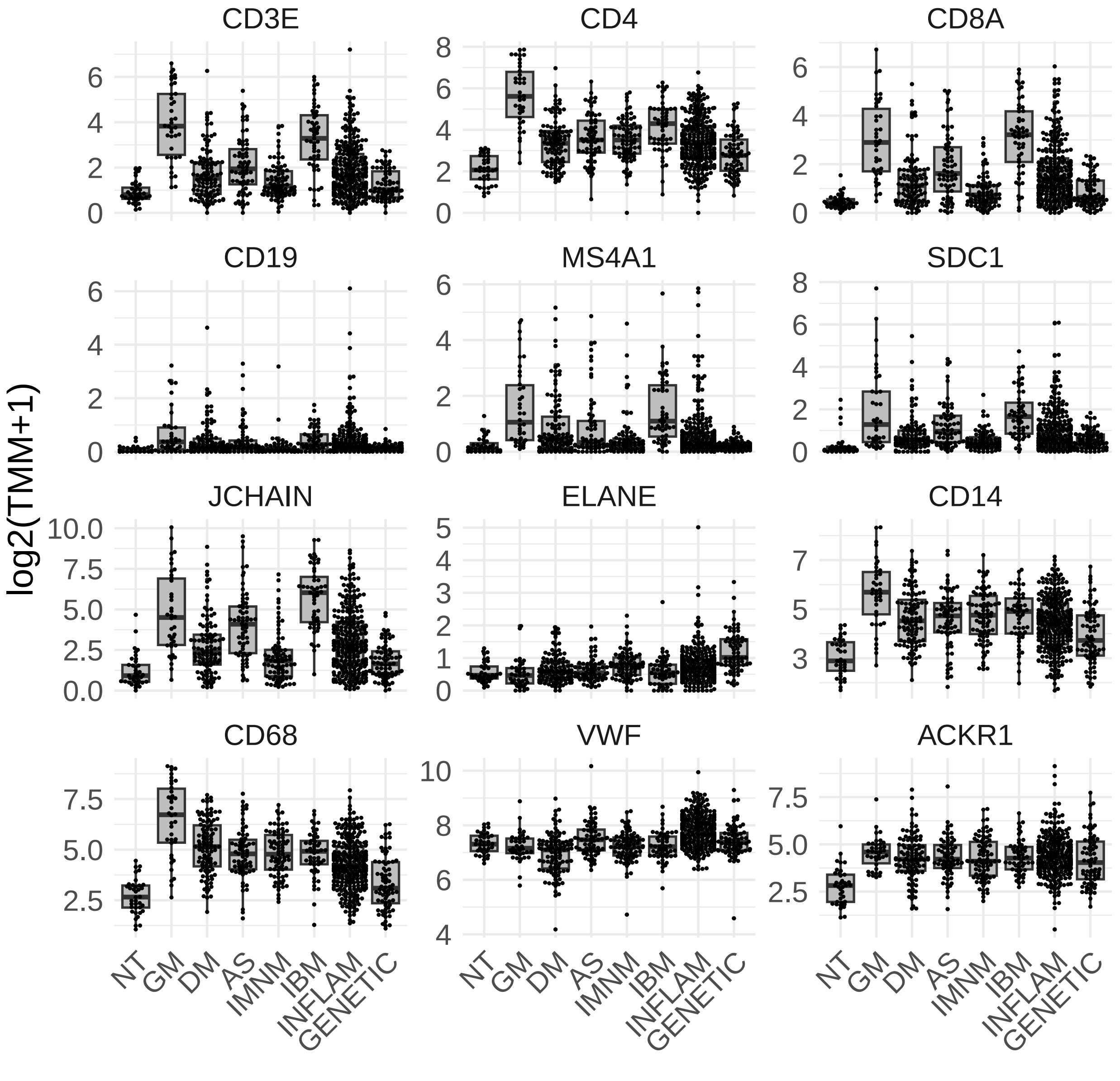

**Supplementary Figure 5.** Expression of transcriptomic markers for HLA and various immunoglobulin isotypes in granulomatous myositis compared to other myopathies. Each dot represents the gene expression value from a single patient. NT: histologically normal muscle biopsies; GM: granulomatous myositis; DM: dermatomyositis; AS: antisynthetase syndrome; IBM: inclusion body myositis; INFLAM: inflammatory myopathies; GENETIC: genetic myopathies.

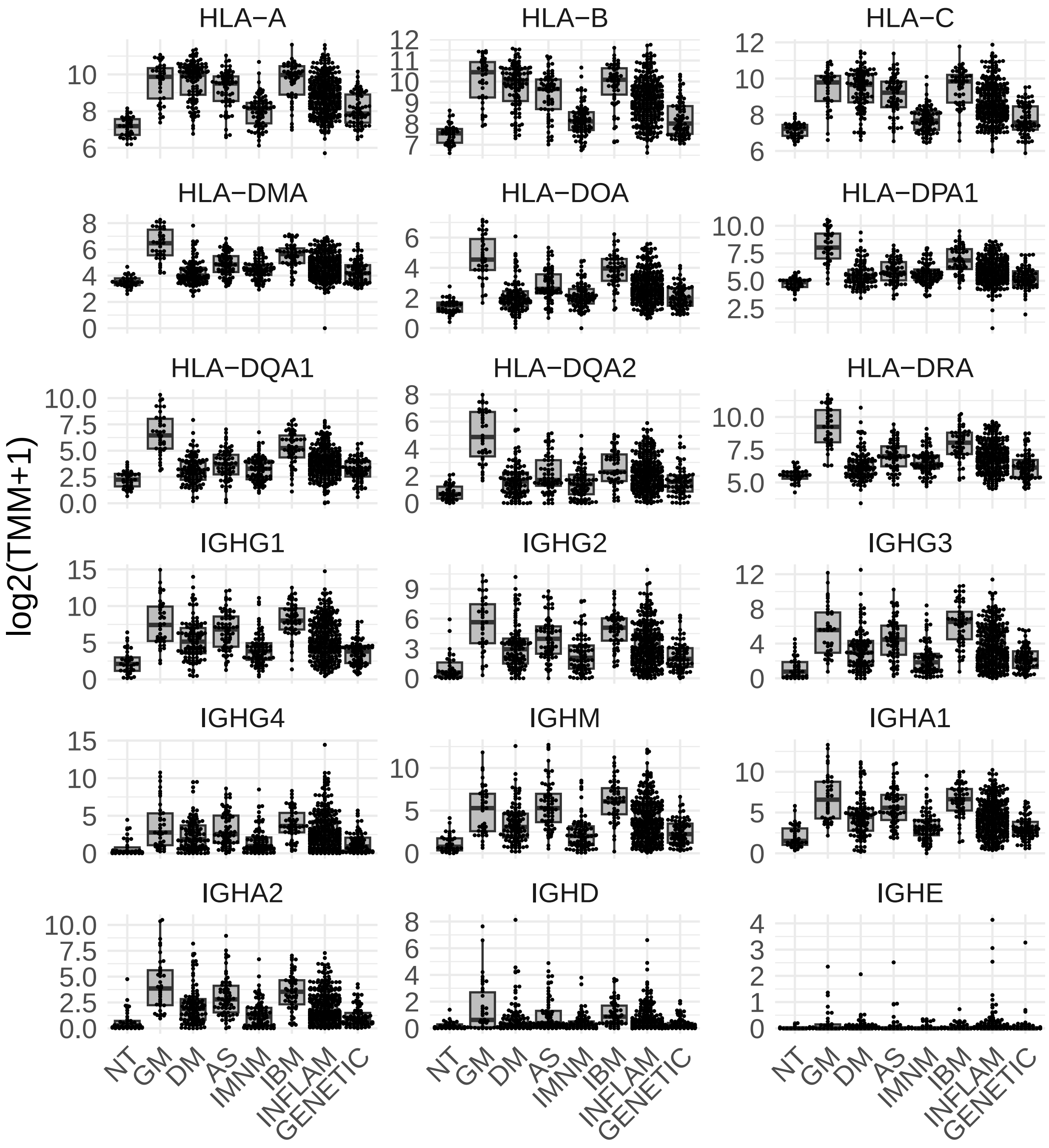

**Supplementary Figure 6.** Expression of transcriptomic markers for mature muscle fibers (MYH7, MYH2, MYH1, ACTA1, TTN) and regenerating muscle fibers (NCAM1, PAX7, MYH3, MYH7) in granulomatous myositis compared to other myopathies. Each dot represents the gene expression value from a single patient. NT: histologically normal muscle biopsies; GM: granulomatous myositis; DM: dermatomyositis; AS: antisynthetase syndrome; IBM: inclusion body myositis; INFLAM: inflammatory myopathies; GENETIC: genetic myopathies.

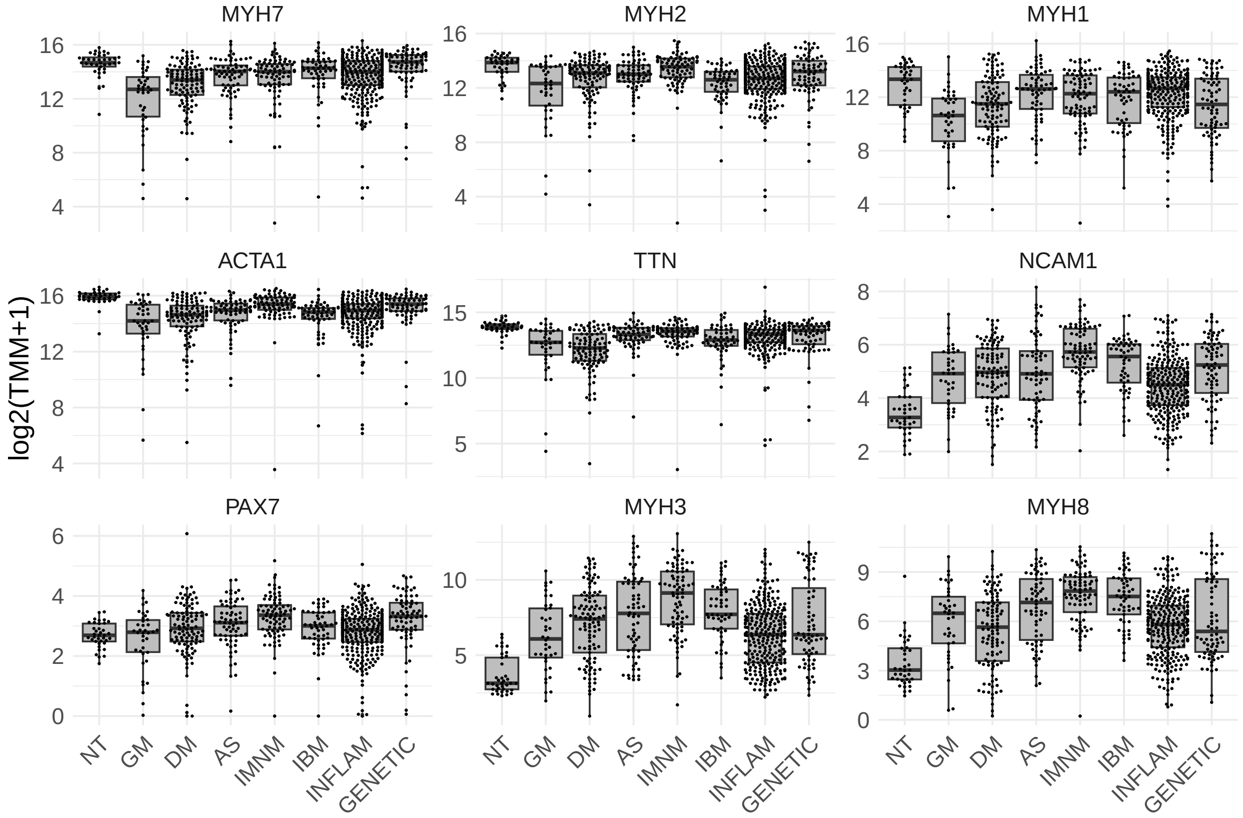

**Supplementary Figure 7.** Expression of mitochondrial transcriptomic markers in granulomatous myositis compared to other myopathies. Each dot represents the gene expression value from a single patient. NT: histologically normal muscle biopsies; GM: granulomatous myositis; DM: dermatomyositis; AS: antisynthetase syndrome; IBM: inclusion body myositis; INFLAM: inflammatory myopathies; GENETIC: genetic myopathies.

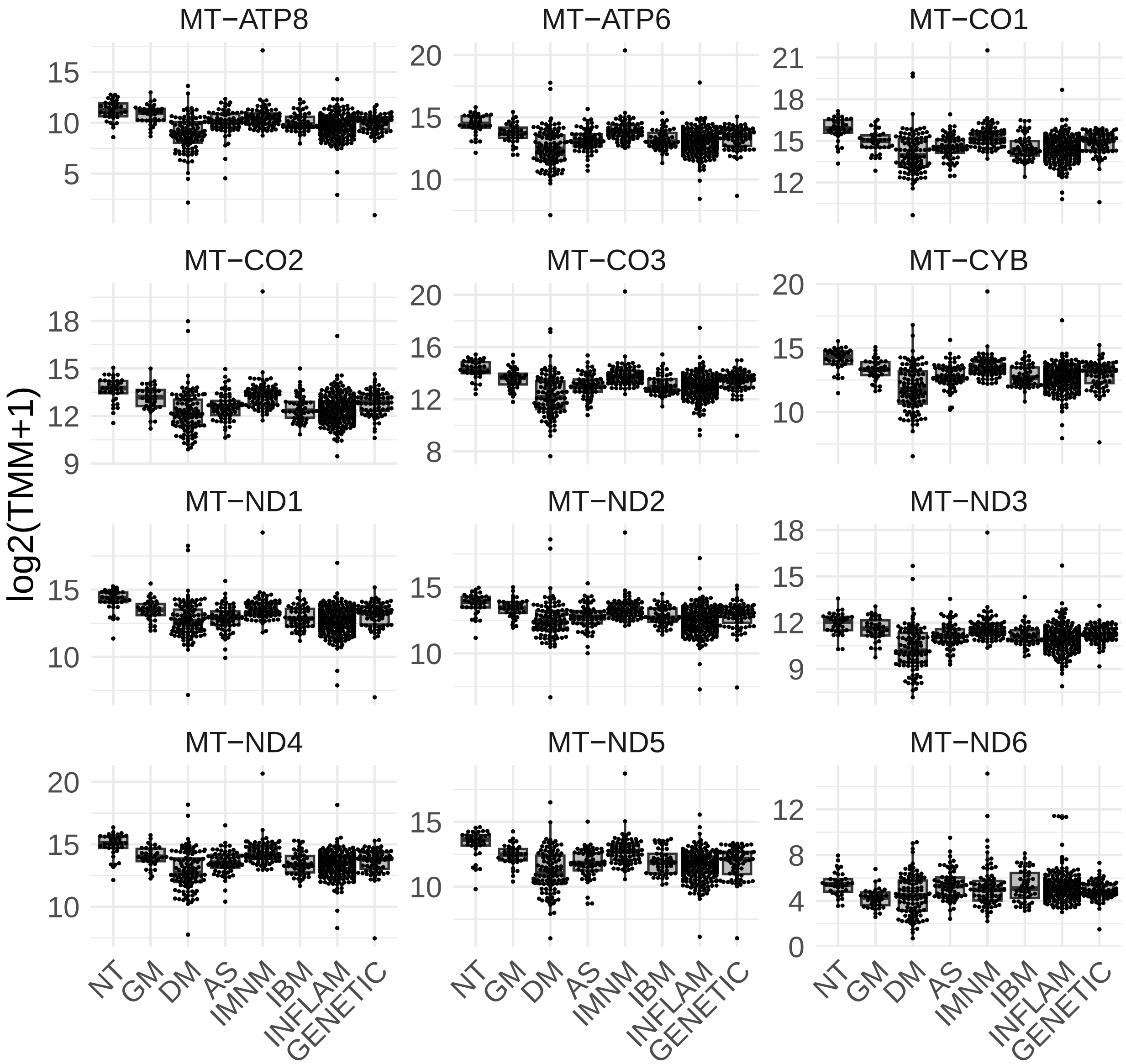

**Supplementary Figure 8.** Expression of significantly different interleukin genes in granulomatous myositis compared to other myopathies. Each dot represents the gene expression value from a single patient. NT: histologically normal muscle biopsies; GM: granulomatous myositis; DM: dermatomyositis; AS: antisynthetase syndrome; IBM: inclusion body myositis; INFLAM: inflammatory myopathies; GENETIC: genetic myopathies.

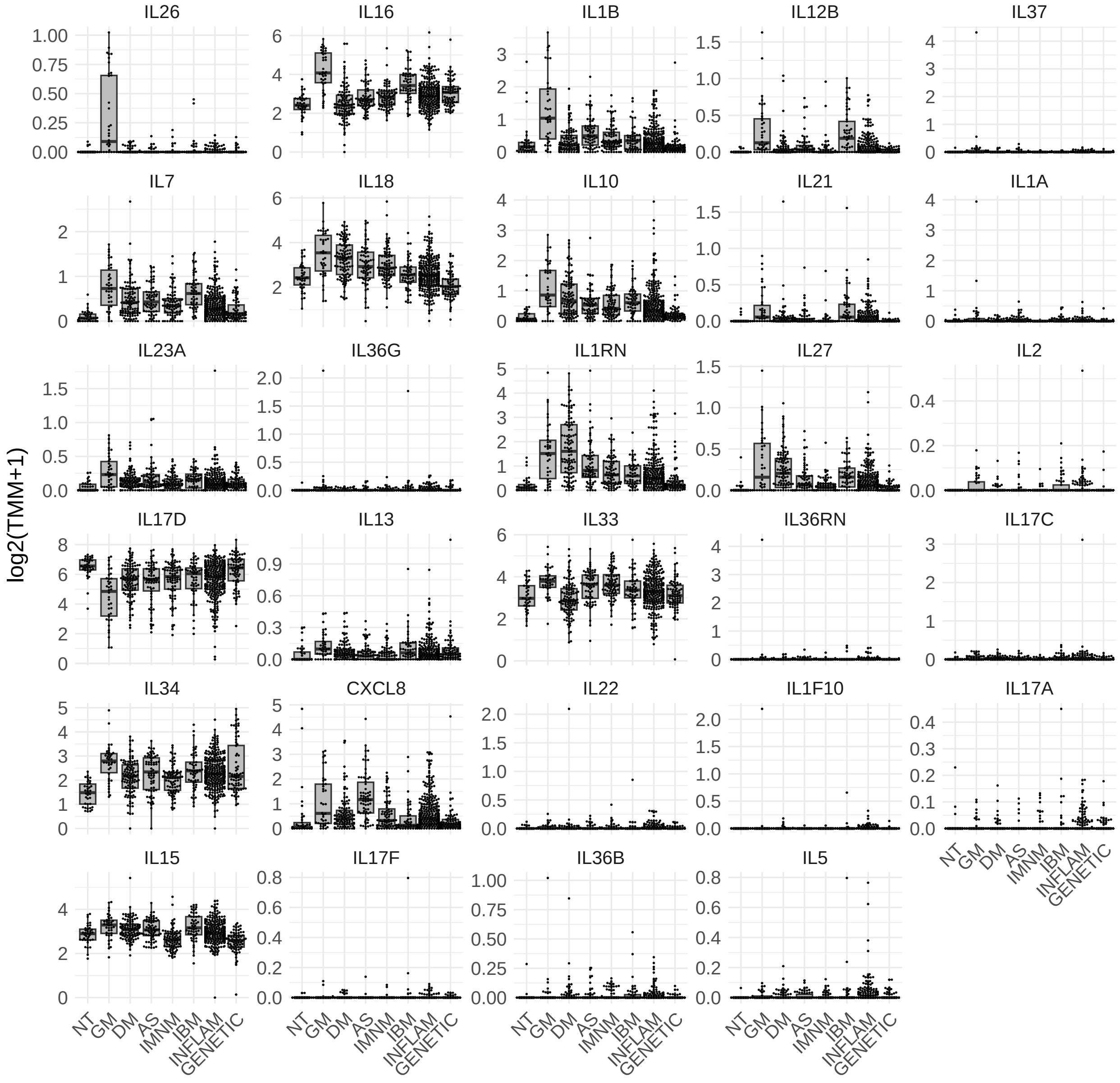

**Supplementary Figure 9.** Expression of significantly different (q-value < 0.05) interleukin receptor genes in granulomatous myositis compared to other myopathies. Each dot represents the gene expression value from a single patient. NT: histologically normal muscle biopsies; GM: granulomatous myositis; DM: dermatomyositis; AS: antisynthetase syndrome; IBM: inclusion body myositis; INFLAM: inflammatory myopathies; GENETIC: genetic myopathies.

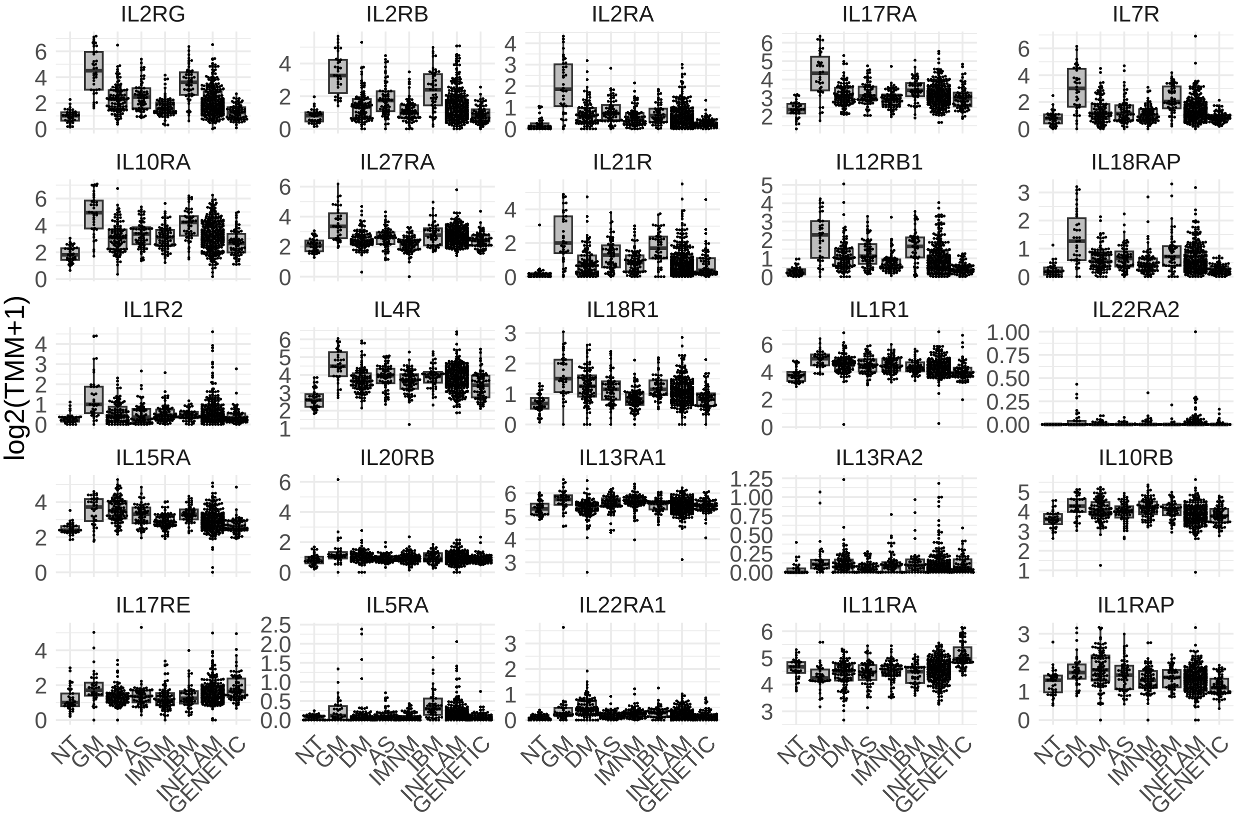

**Supplementary Figure 10.** Expression of checkpoint inhibitor ligands and receptors in granulomatous myositis compared to other myopathies. Each dot represents the gene expression value from a single patient. NT: histologically normal muscle biopsies; GM: granulomatous myositis; DM: dermatomyositis; AS: antisynthetase syndrome; IBM: inclusion body myositis; INFLAM: inflammatory myopathies; GENETIC: genetic myopathies.

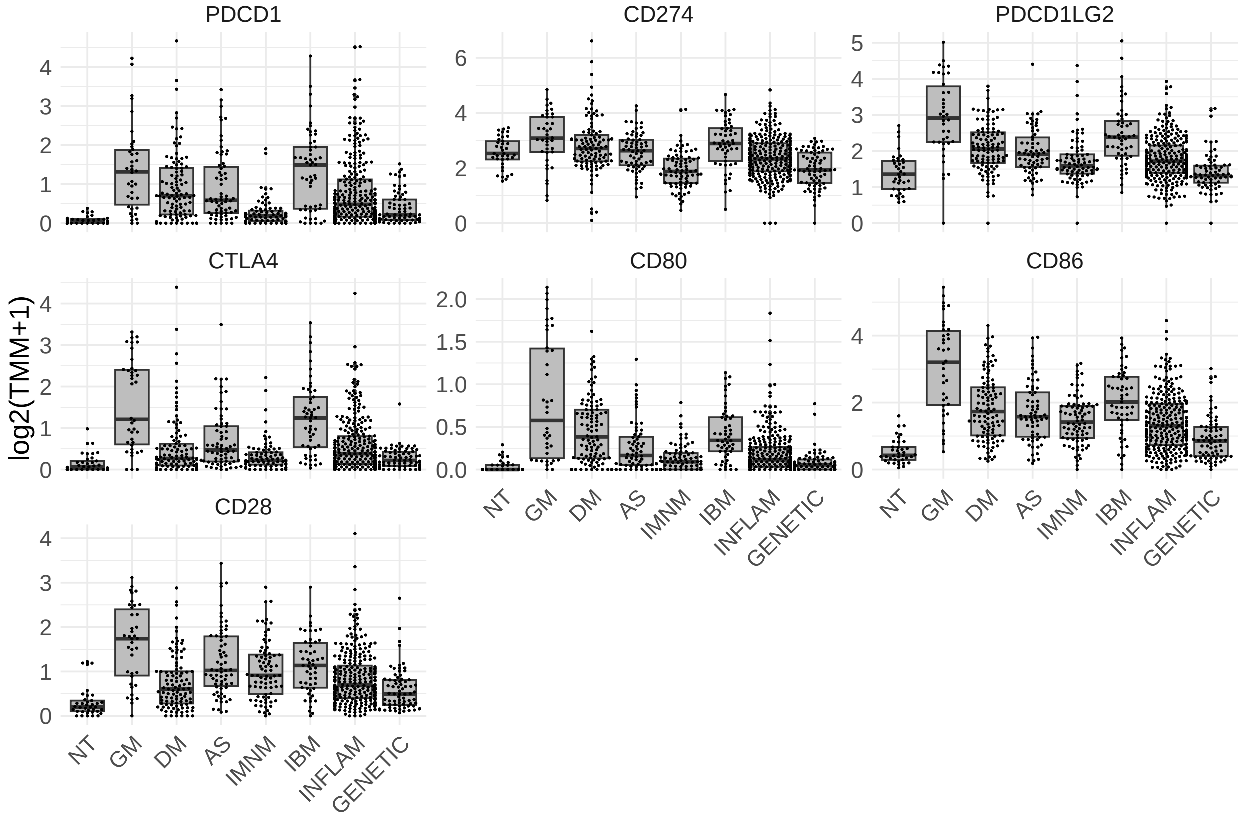

**Supplementary Figure 11.** Expression of significantly different (q-value < 0.05) chemokine genes in granulomatous myositis compared to other myopathies. Each dot represents the gene expression value from a single patient. NT: histologically normal muscle biopsies; GM: granulomatous myositis; DM: dermatomyositis; AS: antisynthetase syndrome; IBM: inclusion body myositis; INFLAM: inflammatory myopathies; GENETIC: genetic myopathies.

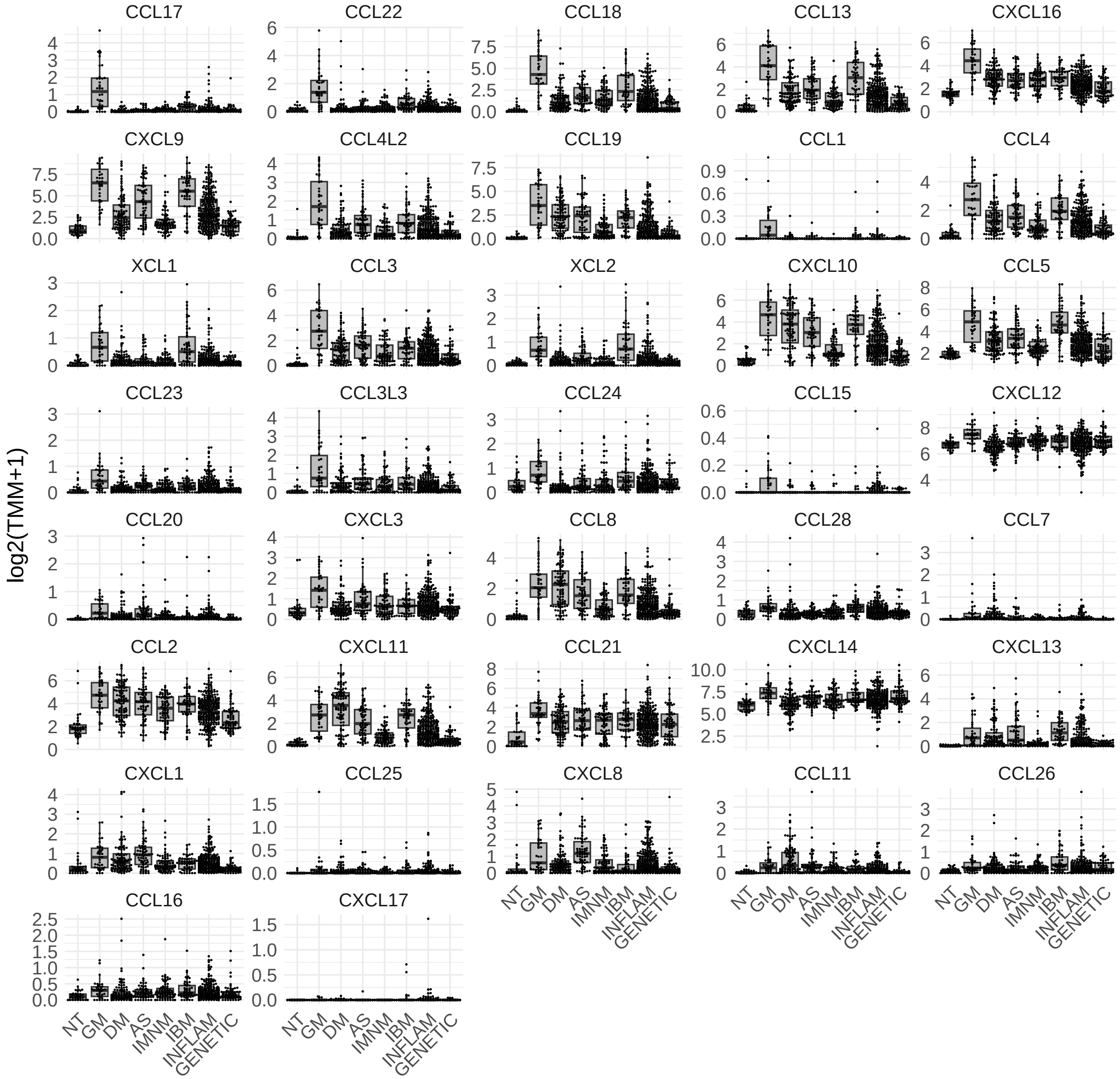

**Supplementary Figure 12.** Expression of significantly different (q-value < 0.05) chemokine receptor genes in granulomatous myositis compared to other myopathies. Each dot represents the gene expression value from a single patient. NT: histologically normal muscle biopsies; GM: granulomatous myositis; DM: dermatomyositis; AS: antisynthetase syndrome; IBM: inclusion body myositis; INFLAM: inflammatory myopathies; GENETIC: genetic myopathies.

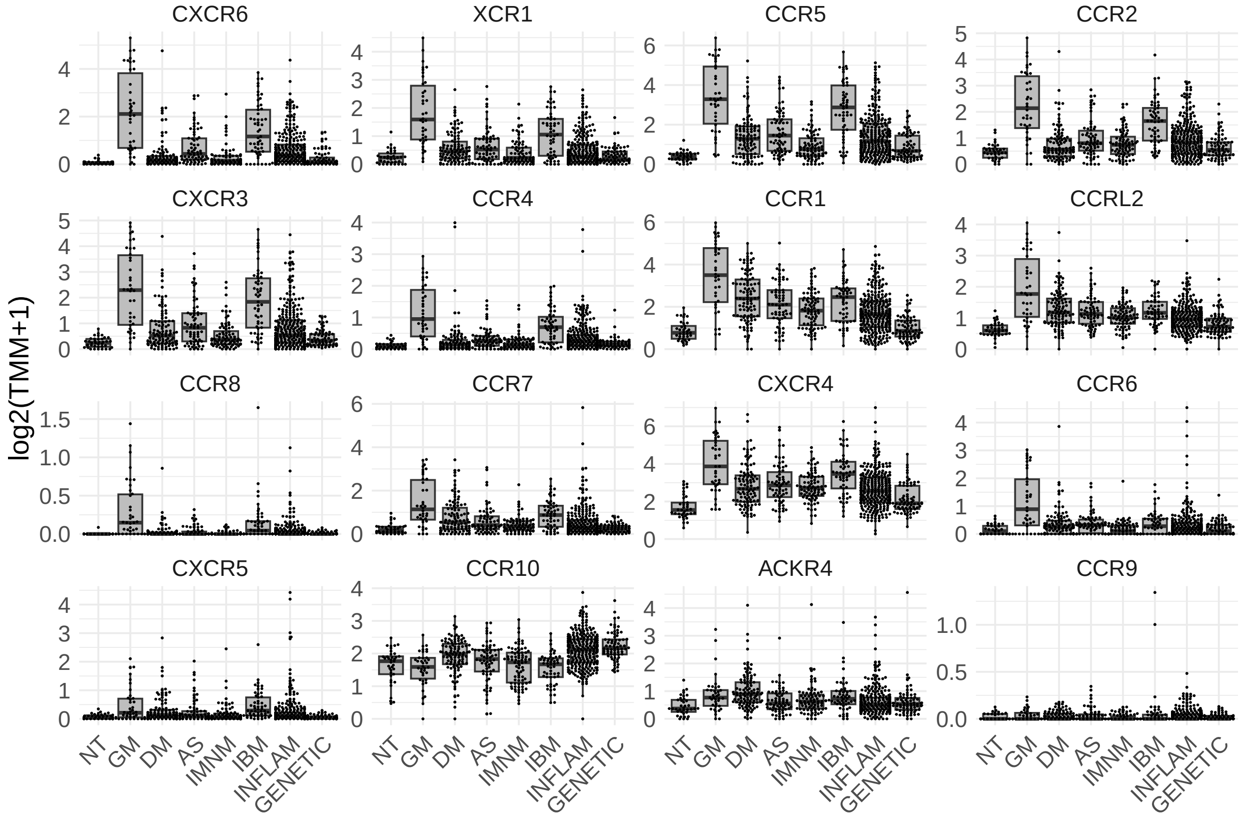

**Supplementary Figure 13.** Expression of significantly different (q-value < 0.05) tumor necrosis factor genes in granulomatous myositis compared to other myopathies. Each dot represents the gene expression value from a single patient. NT: histologically normal muscle biopsies; GM: granulomatous myositis; DM: dermatomyositis; AS: antisynthetase syndrome; IBM: inclusion body myositis; INFLAM: inflammatory myopathies; GENETIC: genetic myopathies.

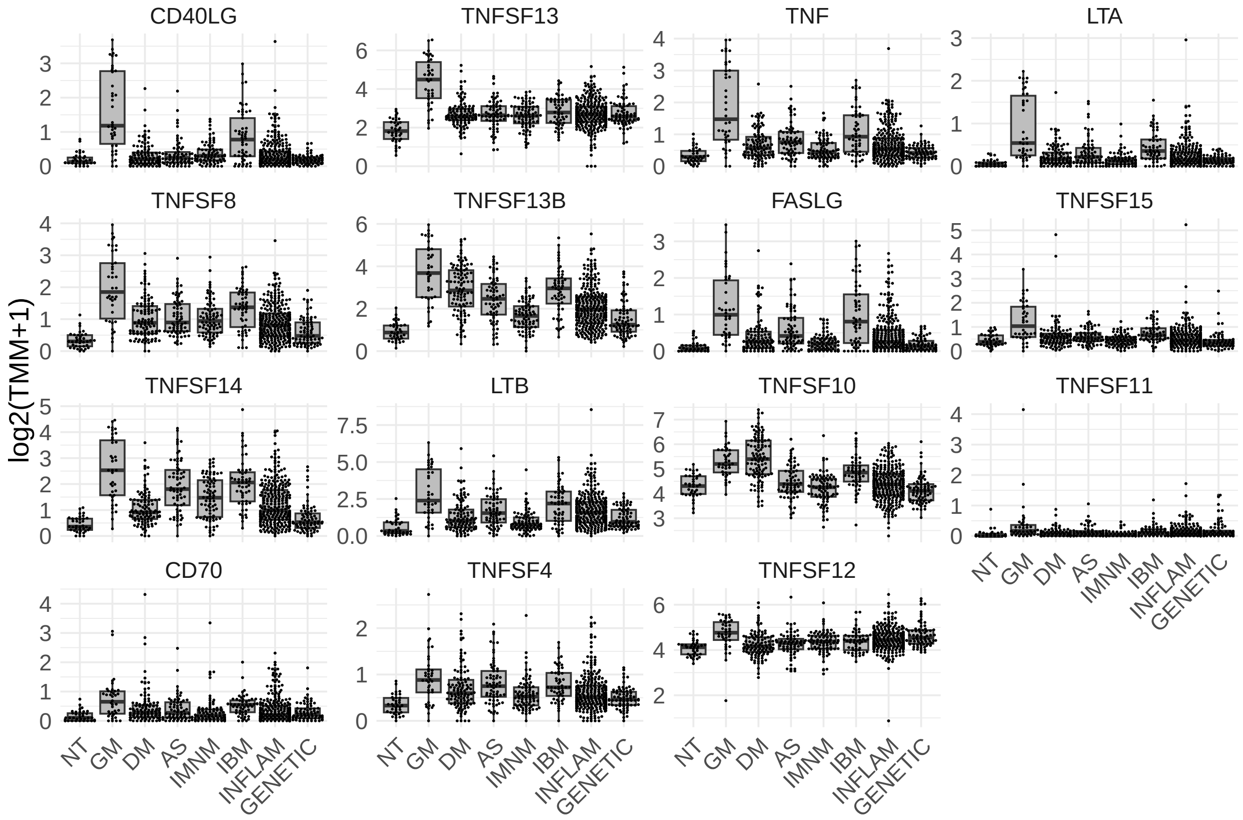

**Supplementary Figure 14.** Expression of significantly different (q-value < 0.05) tumor necrosis factor receptor genes in granulomatous myositis compared to other myopathies. Each dot represents the gene expression value from a single patient. NT: histologically normal muscle biopsies; GM: granulomatous myositis; DM: dermatomyositis; AS: antisynthetase syndrome; IBM: inclusion body myositis; INFLAM: inflammatory myopathies; GENETIC: genetic myopathies.

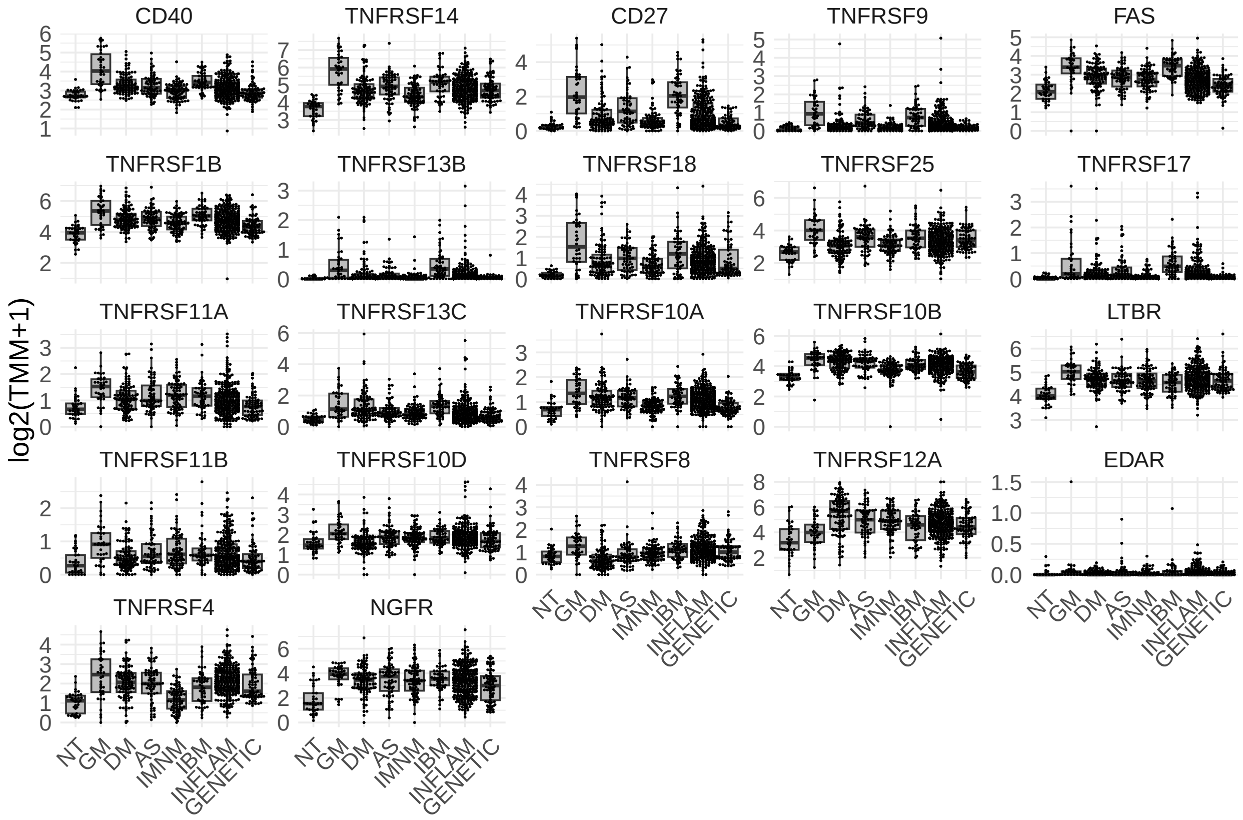

**Supplementary Figure 15.** Expression of significantly different (q-value < 0.05) TGF$\beta$ genes and receptors in granulomatous myositis compared to other myopathies. Each dot represents the gene expression value from a single patient. NT: histologically normal muscle biopsies; GM: granulomatous myositis; DM: dermatomyositis; AS: antisynthetase syndrome; IBM: inclusion body myositis; INFLAM: inflammatory myopathies; GENETIC: genetic myopathies.

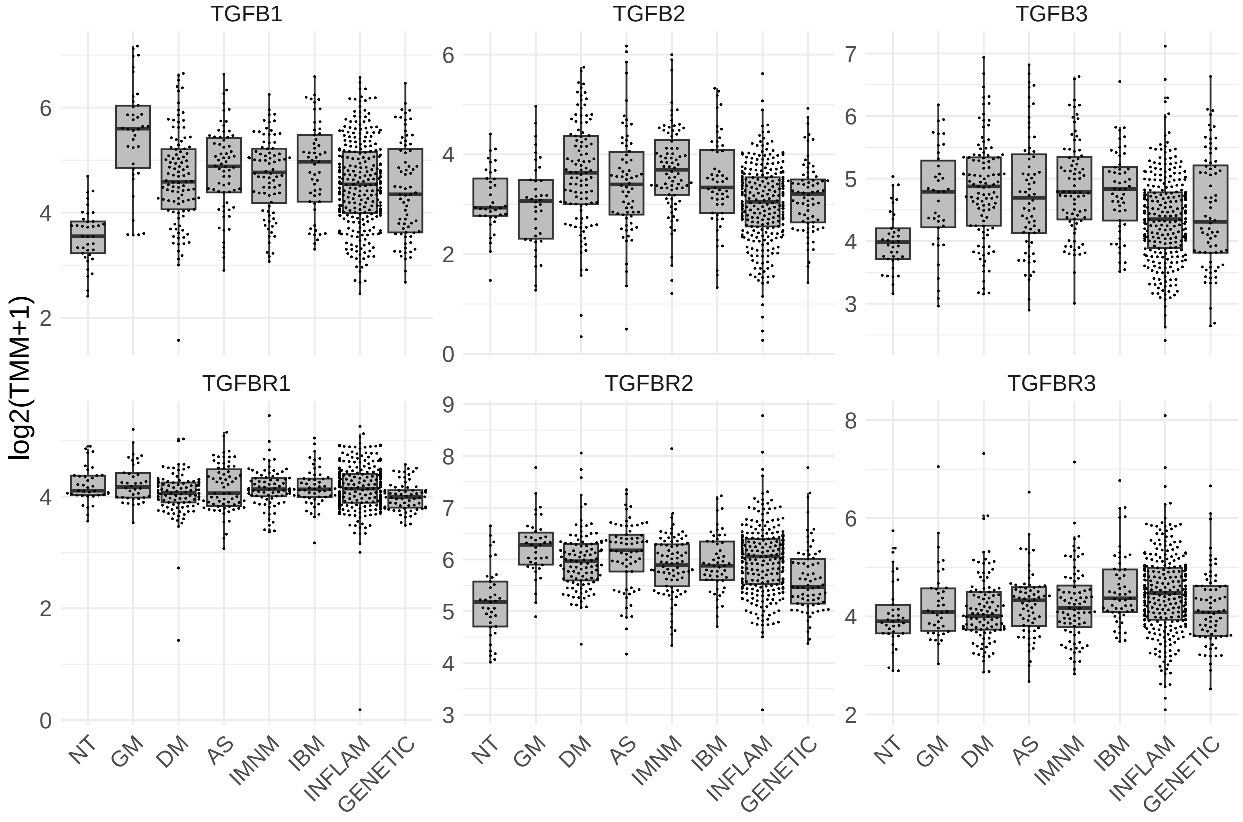

**Supplementary Figure 16.** Distribution of the expression of representative genes in granulomatous myositis.

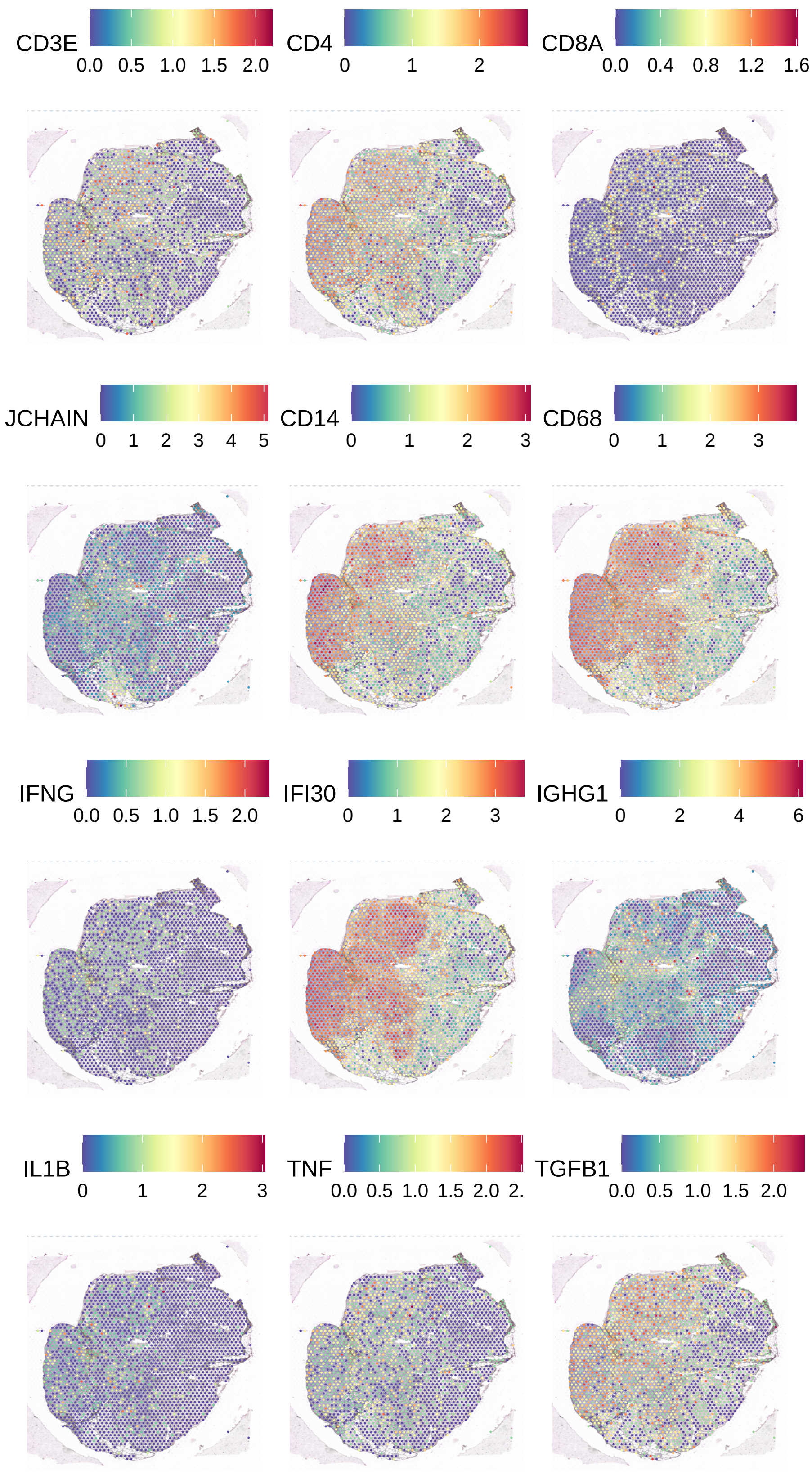

**Supplementary Figure 17.** Expression of top 12 specifically underexpressed genes in granulomatous myositis compared to other myopathies. Each dot represents the gene expression value from a single patient. NT: histologically normal muscle biopsies; GM: granulomatous myositis; DM: dermatomyositis; AS: antisynthetase syndrome; IBM: inclusion body myositis; INFLAM: inflammatory myopathies; GENETIC: genetic myopathies.

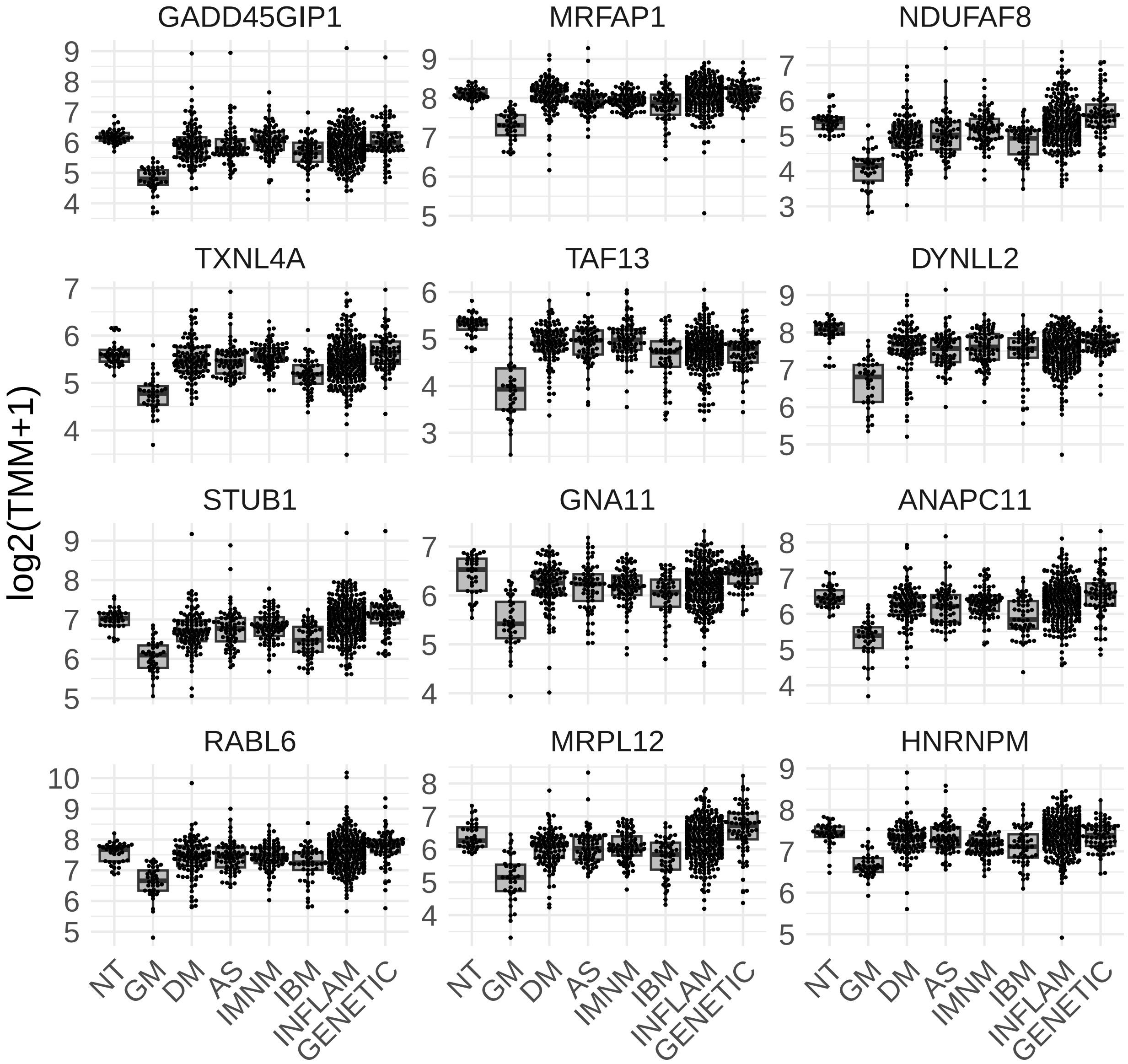

**Supplementary Figure 18.** Correlation of the top 12 specifically underexpressed genes in granulomatous myositis with transcriptomic markers of disease activity, including type 1 interferon-inducible genes (ISG15, MX1), type 2 interferon-inducible genes (GBP2, IFI30), T-cell markers (CD3E, CD4, CD8), macrophage markers (CD14, CD68), markers of muscle differentiation (MYH3, MYH8, NCAM1, PAX7), mitochondrial markers (MT-CO1, MT-CO2), and mature muscle structural proteins (ACTA1, MYH1, MYH2).

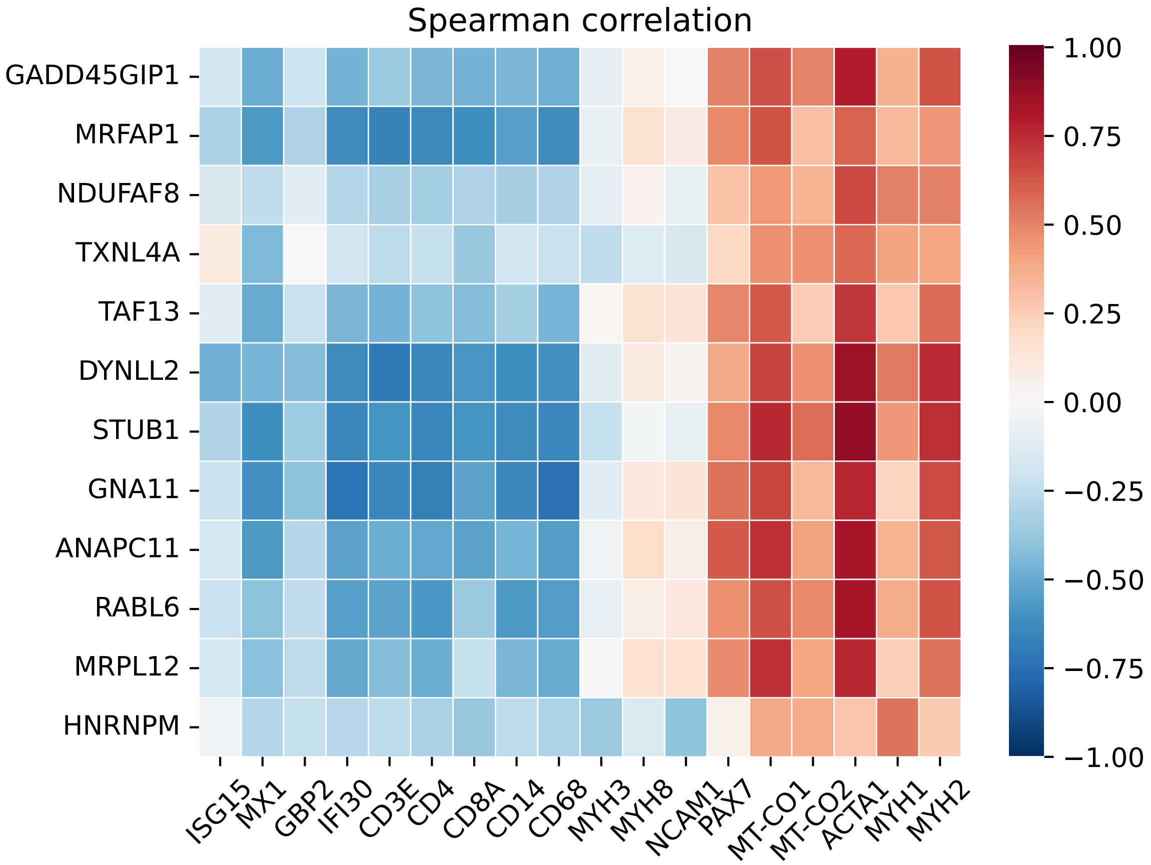

**Supplementary Figure 19.** Expression of top 12 specifically overexpressed genes in granulomatous myositis compared to other myopathies, stratified by cohort location. Each dot represents the gene expression value from a single patient. NT: histologically normal muscle biopsies; GM: granulomatous myositis; DM: dermatomyositis; AS: antisynthetase syndrome; IBM: inclusion body myositis; INFLAM: inflammatory myopathies; GENETIC: genetic myopathies.

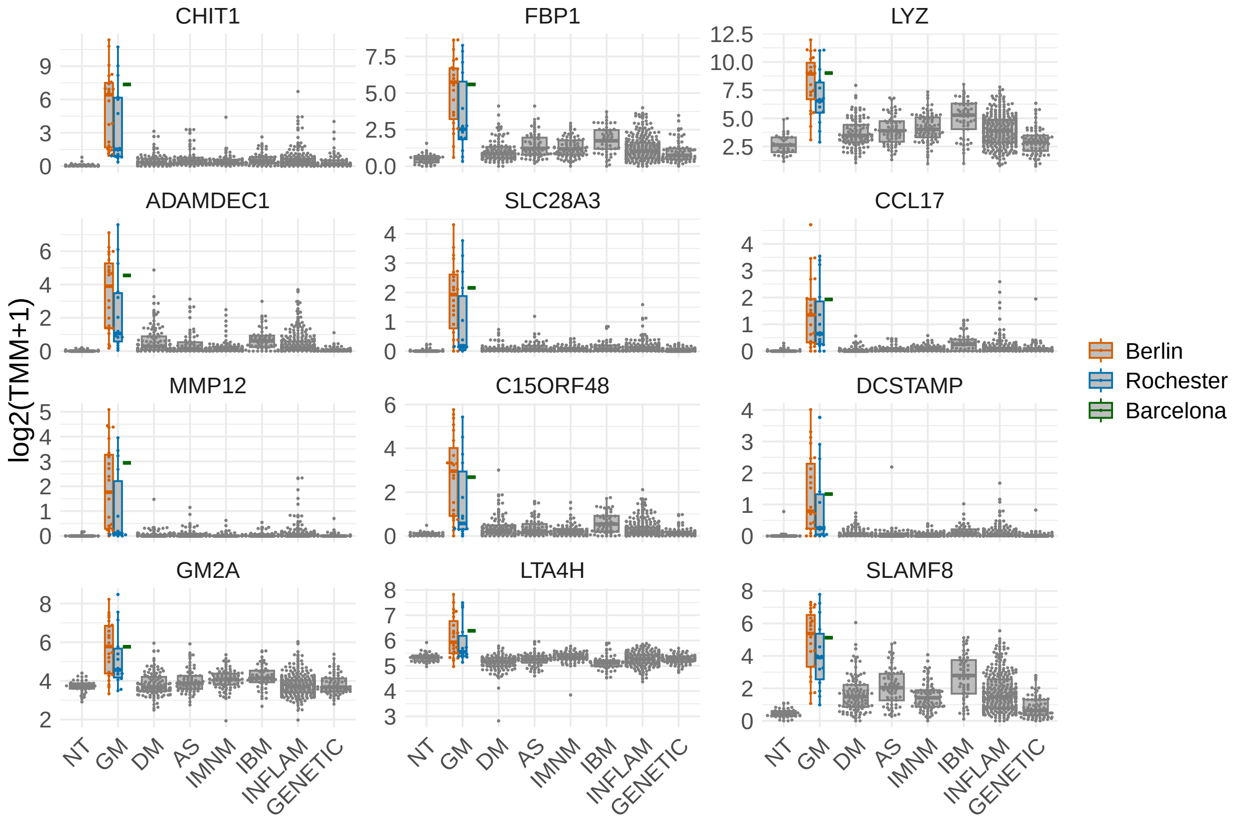
