## Supplementary Tables for "Identification of a distinctive gene signature in granulomatous myositis"

**Supplementary Table 1.** Groups of patients included in the study.

| | N = 722[1] |
|---|---|
| Patient group | |
| Normal biopsies | 37 (5.1%) |
| Granulomatous myositis | 38 (5.3%) |
| Dermatomyositis | 105 (15%) |
| Antisynthetase syndrome | 66 (9.1%) |
| Immune-mediated necrotizing myositis | 80 (11%) |
| Inclusion body myositis | 53 (7.3%) |
| Other inflammatory myopathies | 271 (38%) |
| Genetic myopathies | 72 (10.0%) |
| Patient cohort for granulomatous myositis | |
| Berlin | 23 (61%) |
| Rochester | 14 (37%) |
| Barcelona | 1 (2.6%) |

[1]n (%)

**Supplementary Table 2.** A comparison of machine learning models to classify granulomatous myositis muscle biopsies based on the set of specifically differentially expressed genes identified in the study. Model performance was assessed through stratified 10-fold cross-validation.To construct confidence intervals, we calculated the 2.5% and 97.5% percentiles of performance metrics from 1,000 iterations of the stratified 10-fold cross-validation, each with varying random seeds.

| Model | Accuracy | Area under the ROC curve |
|---|---|---|
| Linear Support Vector Machine (SVM) | 98.6% (98.2%-98.9%) | 99.6% (99.0%-99.9%) |
| Logistic Regression | 98.4% (98.1%-98.9%) | 99.7% (99.4%-99.9%) |
| AdaBoost | 98.0% (97.4%-98.6%) | 99.2% (98.5%-99.7%) |
| Neural Network | 97.7% (97.1%-98.2%) | 96.8% (95.1%-98.2%) |
| Random Forest | 97.2% (97.0%-97.5%) | 97.3% (96.2%-98.1%) |
| Nearest Neighbors | 96.9% (96.5%-97.2%) | 90.5% (88.6%-92.8%) |
| Radial basis function (RBF) SVM | 96.5% (96.3%-96.7%) | 98.3% (97.8%-98.8%) |
| Decision Tree | 95.4% (94.3%-96.5%) | 79.0% (74.3%-84.0%) |
| Gausian Process | 94.7% (94.3%-95.2%) | 50.1% (50.0%-50.1%) |
| Gausian Naive Bayes | 91.7% (91.3%-92.2%) | 87.8% (86.4%-88.8%) |
| Quadratic Discriminant Analysis | 34.6% (29.0%-41.6%) | 58.9% (52.4%-65.2%) |
